# Multi-ancestral GWAS with the VA Million Veteran Program enables functional interpretation of rheumatoid arthritis alleles

**DOI:** 10.64898/2026.04.22.26351423

**Authors:** Saori Sakaue, Doris Yang, Harrison Zhang, Daniel Posner, Zachary Rodriguez, Zoe Love, Jing Cui, Ashley Budu-Aggrey, Yuk-Lam Ho, Lauren Costa, Paul Monach, Sicong Huang, Kazuyoshi Ishigaki, Connor Melley, Vidisha Tanukonda, Rahul Sangar, Monika Maripuri, Sara Morini Sweet, Vidul Panickan, Gregory McDermott, Jennifer S. Hanberg, Thomas Riley, Vincent Laufer, Yukinori Okada, Ian Scott, S. Louis Bridges, Joshua Baker, VA Million Veteran Program, Peter W. Wilson, J. Michael Gaziano, Chuan Hong, Anurag Verma, Kelly Cho, Jennifer E. Huffman, Tianxi Cai, Soumya Raychaudhuri, Katherine P. Liao

## Abstract

Rheumatoid arthritis (RA) is a heritable and common autoimmune condition. To date, most genetic associations were derived from individuals with either European or East Asian ancestries. Here, we applied a multimodal automated phenotyping strategy to define RA and performed a genome-wide association study (GWAS) of RA in the Million Veteran Program (MVP), including underrepresented African American (AFR) and Admixed American (AMR) populations. Meta-analyses with previous RA cohorts identified 152 autosomal genome-wide significant loci, of which 31 were novel. Inclusion of multi-ancestry data dramatically improved fine-mapping resolution. Functional characterization of these loci using single-cell transcriptomic and chromatin data suggested new RA genes such as *CHD7* and *CD247*. We identified underappreciated functional roles of fine-grained immune cell states other than T cells, such as B cell and myeloid cell states. We observed that multi-ancestry polygenic risk scores using our data demonstrated better predictive ability, especially for AFR and AMR populations.

## Main

RA is one of the most common autoimmune diseases, affecting approximately 1% of individuals worldwide^1^ and is without a cure. RA has strong heritable components with an estimated heritability of ∼60% in twin studies^2^. Previous RA GWAS have identified >120 genome-wide significant loci to date^3^. These alleles have provided key insights into autoimmune mechanisms, including gene regulatory effects in T cells^4^ and roles in antigen recognition and presentation by HLA^5,6^. RA susceptibility alleles may point to genes with therapeutic potential^7^.

However, prior genetic studies have relied predominantly on data from European (EUR) and East Asian (EAS) populations, which comprised 85.5% and 13.7%, respectively, in the most recent multi-ancestral study^3^; AFR and AMR populations remain severely underrepresented, comprising only 0.52% and 0.043%, respectively, consistent with broader patterns in the field^8,9^. Epidemiological studies suggest that RA patients of AFR and AMR ancestry may experience severe disease, perhaps due to health disparities, such as delayed treatment^10–12^. Elucidating the genetic basis of RA in underrepresented populations is expected to (1) improve generalizability across populations, (2) increase power for novel locus discovery, (3) refine fine-mapping by leveraging diverse linkage disequilibrium (LD) structures^13^, and (4) improve genetic risk prediction in understudied populations^14^.

Large-scale biobank data offer an opportunity to address this gap. The Department of Veteran Affairs (VA) Million Veteran Program (MVP)^15^, a longitudinal health, genomic, and precision medicine cohort, provides both population diversity and deep phenotypic data from electronic health records (EHR). AFR and AMR representation in MVP is among the largest across genomic biobanks^15–18^. It is challenging in biobanks to define RA patients, since the disease is relatively uncommon, and diagnostic codes and patient self-reports can be inaccurate. In this context, we applied a multimodal automated phenotyping strategy to improve the accuracy of RA classification and performed an updated RA GWAS by integrating MVP biobank data with 37 international RA cohorts (Rheumatoid Arthritis Consortium International (RACI); **Figure 1a** and **1b**). This increased statistical power identified 31 novel loci. The multi-ancestry nature of this study enhanced statistical fine-mapping and prioritization of putative causal variants. Functional genomics analyses using single-cell multiomics of RA synovial tissue identified target genes in disease-relevant cell types. These results provide new insight into the genetic architecture of RA and guide functional follow-up studies.

**Figure 1.**
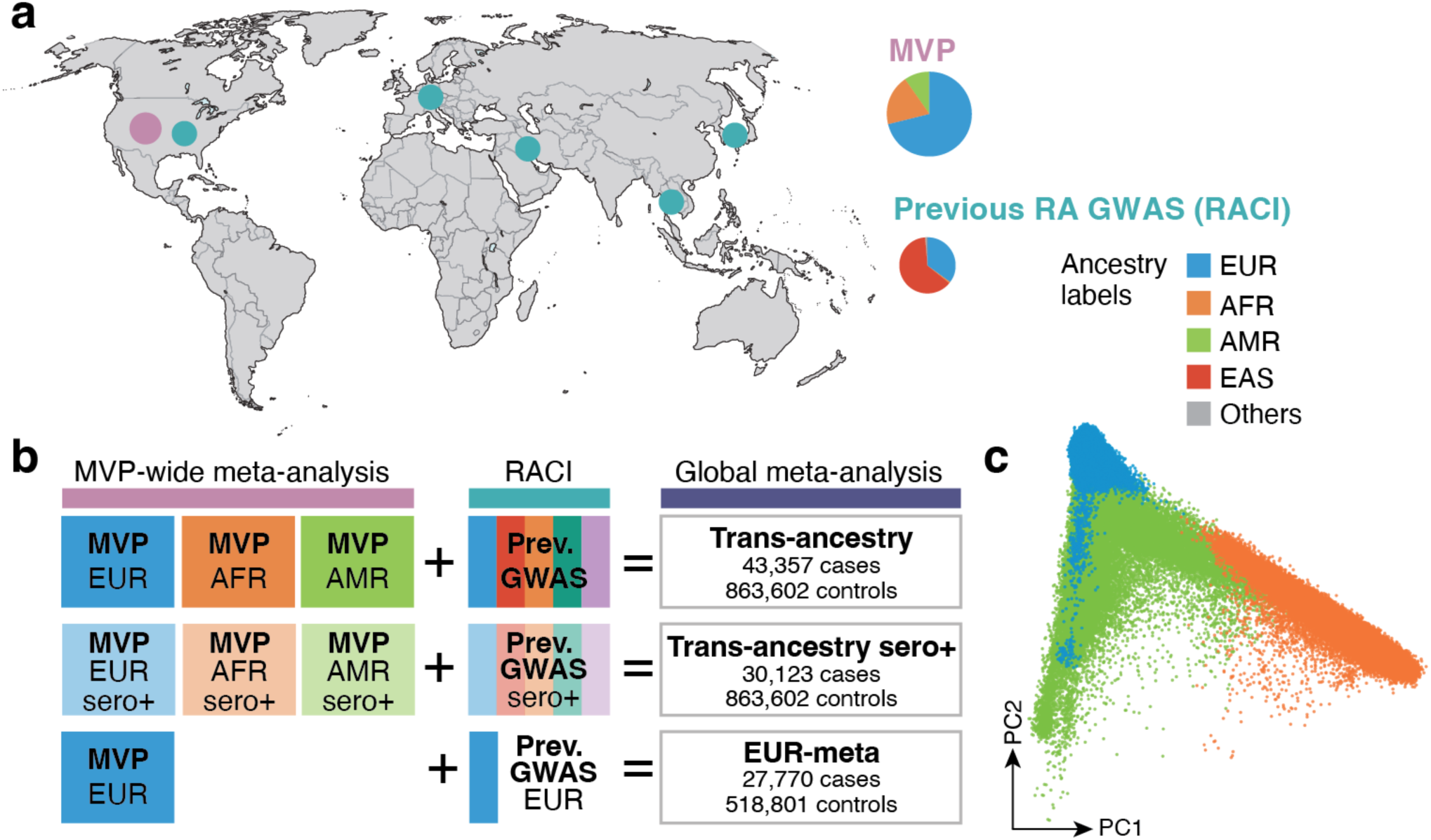
Overview of our study. **a.** The geographic and ancestral characteristics of our RA GWAS. Circles in the map reflect the sample size of the cohorts, colored in pink for the MVP cohort and in green for previous GWAS cohorts of RACI. Pie charts represent population representation in each of GWAS. **b**. Graphic overview of meta-analyses strategies and sample sizes. **c**. Genetic PCA of individuals at MVP projected onto 1KG samples, colored by assigned population labels.

## Results

The MVP is a longitudinal biobank linked with clinical EHR data, which began enrollment in 2011 and reached its one-millionth Veteran in 2023^15,19^. We conducted a primary GWAS involving 7,486 RA cases and 623,453 controls without RA in the MVP Genomics Release 4. We included 121,316 AFR (1,397 cases), 60,225 AMR (669 cases) and 449,398 EUR (5,420 cases) samples (**Figure 1c**; **Supplementary Table 1**). The mean age of participants was 61.7 years and 91.2% were male, reflecting the Veteran population (**Supplementary Table 1**). The prevalence of RA in MVP was 1.2% in AFR, 1.1% in AMR, and 1.2% in EUR, consistent with previous population estimates ^20,21^. The primary GWAS and cross-population meta-analyses in MVP identified nine autosomal genome-wide significant loci outside the major histocompatibility complex (MHC) region (**Figure 2a**, **Supplementary Figure 1a–c**), including one novel locus at *PTPN11* (**Supplementary Table 2**). We observed little evidence of population structure, with well-calibrated genomic inflation^22^ (1.026) and LDSC intercept ^23^ (1.01) (**Supplementary Table 3**).

**Figure 2.**
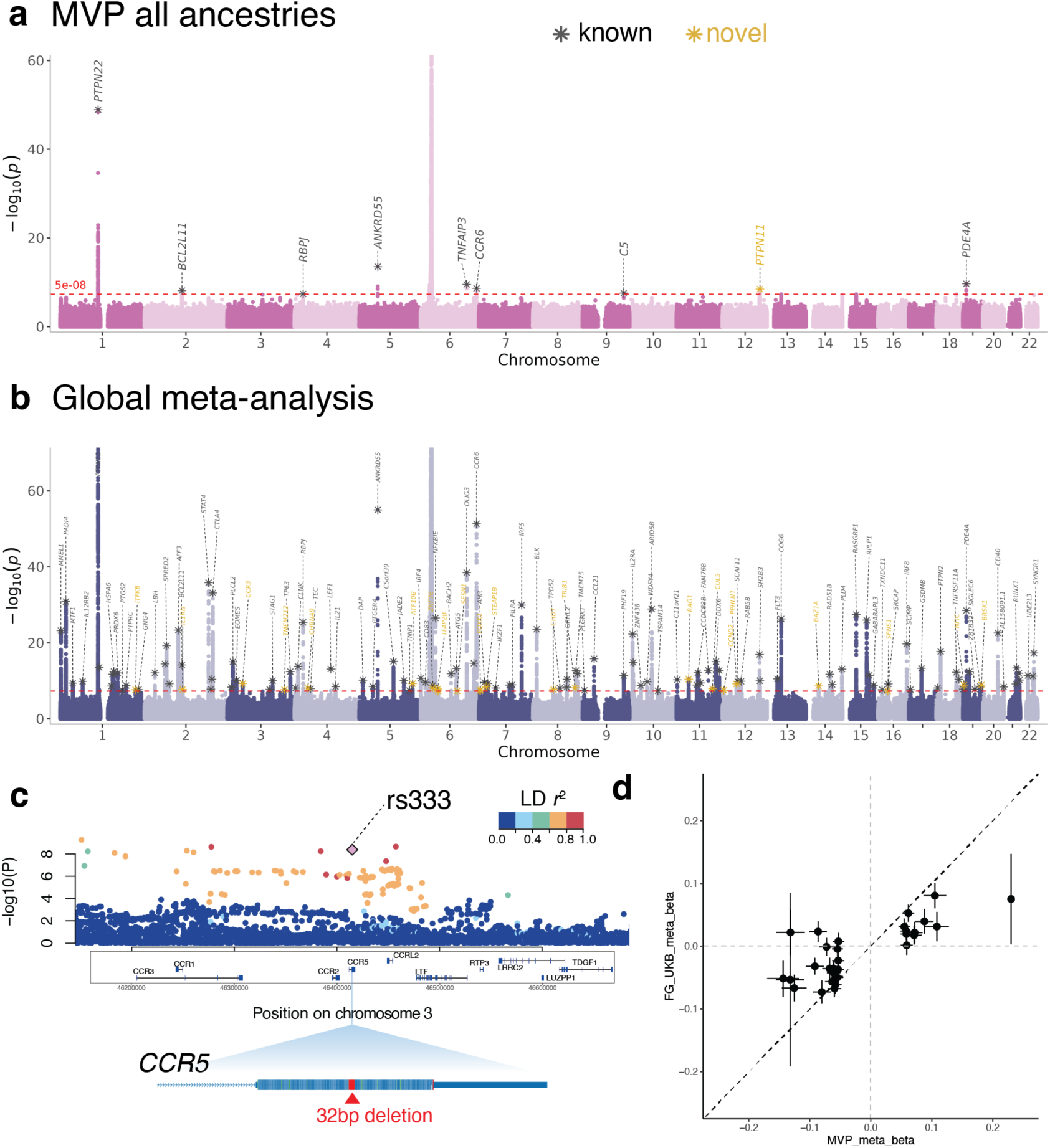
Cross-ancestry meta-analyses of RA. **a.** A Manhattan plot of MVP RA GWAS across AFR, AMR and EUR ancestries. Lead variants with asterisks are annotated by the closest genes and colored in grey (previously reported loci) or in yellow (novel loci). **b**. A Manhattan plot of joint meta-analyses between MVP and all previous RACI cohorts. **c**. A regional plot for *CCR5* loci. Colors represent the LD *R*^2^ values from the lead variant (rs333) as shown in the color scale. The bottom plot shows the location of the exonic deletion. **d**. Concordance of effect sizes (beta) from our novel loci between our study (x-axis) and meta-analysis of FinnGen and UK Biobank (y-axis). Bars indicate standard errors.

One challenge in biobank genomic studies is accurate phenotyping. Population-based biobanks, such as UK Biobank^16^, often use billing codes such as international classification of disease (ICD) codes to assign medical events, but ICD-based phenotyping alone can lead to misclassification and varying accuracy^24,25^. In RA, misclassification with low positive predictive value has been reported^26^. Therefore, we applied a machine learning approach, knowledge-driven online multimodal automated phenotyping (KOMAP), incorporating both coded electronic health records (EHR) data and information extracted from clinical notes with natural language processing^27^. In cross-validation, KOMAP achieved a positive predictive value of 0.93 compared to 0.64 using ≥2 RA ICD codes (**Methods**). Classifying RA with KOMAP increased the number of genome-wide significant loci (P<5×10⁻⁸; 9 vs. 8) and strengthened effect sizes of 109 known autosomal RA risk alleles (paired *t* test t=7.2, two-sided *P*=7.0×10^−11^; **Supplementary Figure 2**) compared to the ICD code definition. This is consistent with the idea that a GWAS with ICD may include misclassified cases and controls.

We then conducted a cross-cohort meta-analyses of RA cohorts from RACI^3^ with MVP. We additionally conducted a meta-analysis restricting cases to seropositive RA (i.e., positive either for RF or anti-CCP antibody), given their distinct phenotypic and genetic architecture compared with seronegative disease (**Supplementary Table 1**). We identified 152 genome-wide significant loci (excluding the MHC region), of which 31 were novel (**Figure 2a**; **Supplementary Figure 3a-c**; **Supplementary Table 4**). Among the novel loci was a frameshift 32-bp deletion (‘Δ32 mutation’) and high-confidence loss-of-function variant^28,29^ rs333 in *CCR5* (beta = −0.11, *P*=4.2×10⁻⁹), a chemokine receptor involved in lymphocyte migration to inflammatory and immune-surveillance sites^30^ (**Figure 2c**). This variant is under strong selection^31^ and was reported for its association with HIV resistance^32,33^ and recently with plasma cytokine CCL4 levels.^34^ We estimated RA SNP-heritability in EUR to be 0.078 (SE: 0.0059) using stratified-linkage disequilibrium score regression (S-LDSC)^3,4^.

To assess replication of novel loci, we used GWAS summary statistics from two large biobanks, UK Biobank^16^ and FinnGen^35^ phenotyped using ICD codes for RA (344 cases and 420,187 controls in UK Biobank and 11,918 cases and 488,425 controls in FinnGen); 28 of the 31 novel loci had directional concordance (effect size Pearson’s correlation *r*=0.78) with 16 reaching nominal significance (P<0.05) with 100% directional concordance (**Figure 2d**; **Supplementary Figure 4**). Absolute effect sizes were systematically larger in MVP than in FinnGen or UK Biobank (0.082 vs. 0.038; paired *t*-test *P*=1.2×10^−7^), possibly reflecting improved phenotyping in MVP. We noted that in these biobanks, there may be limited power due to inaccurate phenotyping and a relatively small number of RA cases.

To define candidate causal variants in genome-wide significant loci, we performed statistical fine-mapping of 152 loci using the approximate Bayes factor (ABF) method^36^. 60 loci had 95% credible sets (CS) with <10 variants, including 10 loci with a single variant (**Figure 3a**). Among 114 previously reported loci from a trans-ancestry study, we observed significantly smaller 95% CS sizes in our GWAS (paired Wilcoxon signed-rank test *P*=9.7×10⁻⁵) and higher posterior inclusion probabilities (PIP) for lead variants (paired t test *P*=2.9×10⁻³; **Figure 3b**), showcasing increased power and diverse LD structures across ancestries.

**Figure 3.**
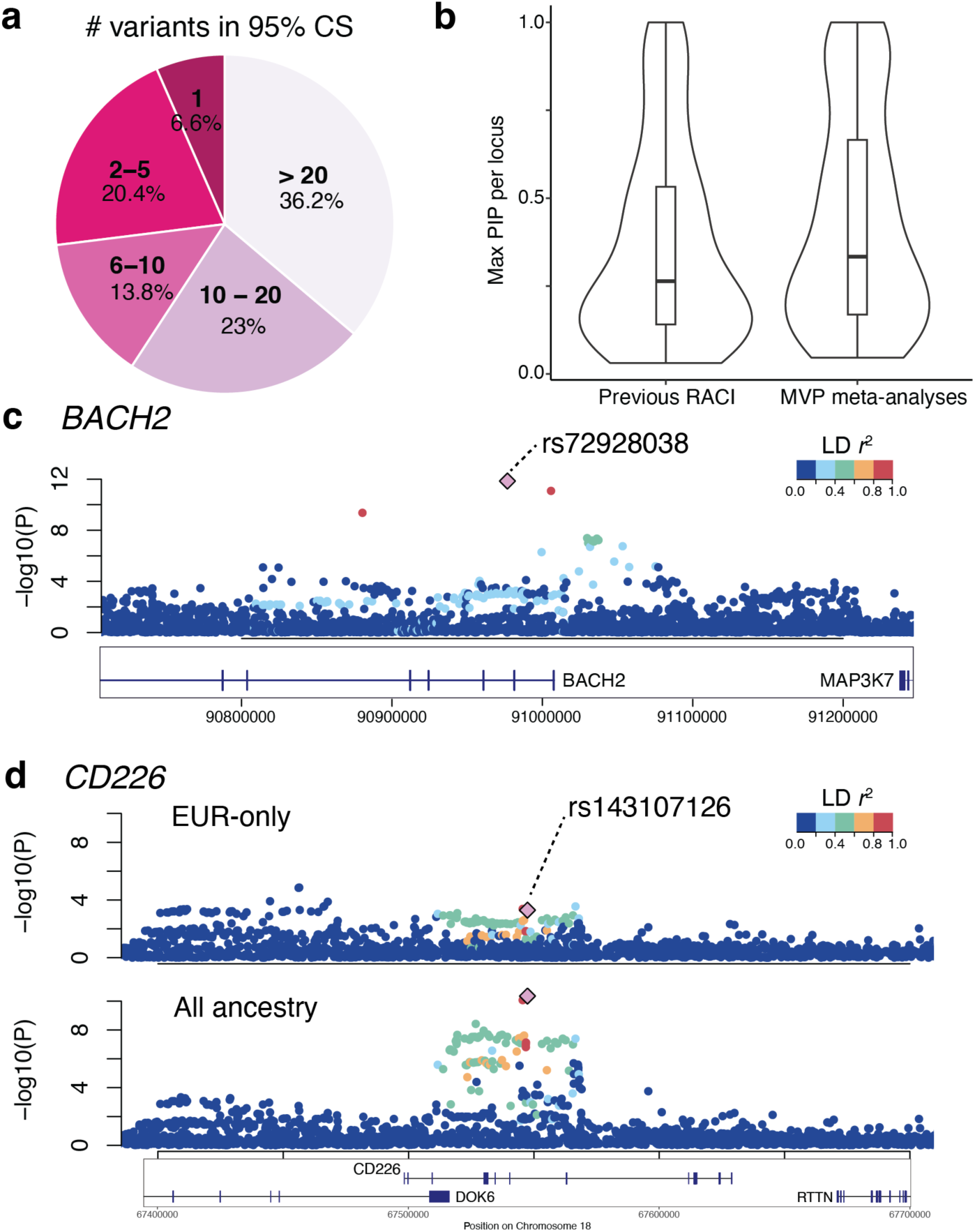
Statistical fine-mapping of RA loci. **a.** The number of fine-mapped variants in 95% credible set (CS) of genome-wide significant loci in our RA GWAS. **b**. Comparison of maximum fine-mapping PIP for each previously reported locus between previous GWAS and our meta-analyses including the MVP. **c**. Previously reported BACH2 locus with a highly probable causal intronic variant rs72928038 (PIP = 0.84). **d**. A deletion rs143107126 at CD226 identified as a causal variant with PIP = 0.63 in all ancestry analyses (bottom) while EUR only analyses had much lower PIP.

For example, at *BACH*2, a well characterized transcription factor in regulatory T cell (T_reg_) and B cell differentiation, we identified a single high-confidence variant, rs72928038 (PIP=0.84; previously only 0.35) ^37,38^ within an intronic enhancer functionally active in T cells (**Figure 3c**). 103 trans-ancestry loci (75%) had fewer 95% CS variants and higher maximum PIP than observed in the EUR-only analyses. As an example, rs143107126 was identified as the most probable causal deletion (PIP = 0.63) at an intronic region of *CD226*, whereas the EUR-only meta-analysis contained 4,424 variants in the 95% CS with much lower PIP (**Figure 3d**). 15 of the 31 new loci had higher minor allele frequencies in AFR or AMR than EUR. For example, we identified rs4265093 at the novel *STEAP1B* locus (PIP = 0.38; 8 variants in the 95% CS), which is more common in AFR and AMR (MAF = 0.17 and 0.16) than in EUR (MAF = 0.098). In summary, the diverse ancestries available in this GWAS resolved LD structure and refined putative causal variants.

142 RA loci (93%) did not include a coding variant within the credible sets, suggesting non-coding variants may drive RA risk. To define their functional variants, effector genes, and cell-type context, we integrated data from single-cell functional genomics data.

First, we determined disease-relevant cell types by testing enrichment of fine-mapped variants in cell-type-specific regulatory elements from single-cell ATAC-seq of synovial tissues from RA and osteoarthritis (OA) patients^39^. Using g-chromVAR^40^, RA variants were most enriched in accessible chromatin of CD4 Tfh/Tph/Treg cells (*P*=2.8×10⁻⁴), CD8 T cells (*P*=2.8×10⁻⁴), and CD4 memory cells (*P*=7.3×10⁻⁴) in inflamed synovium (**Figure 4a**). Tph CD4⁺ T cells are known to play a pathogenic role in RA synovial inflammation^41^.

**Figure 4.**
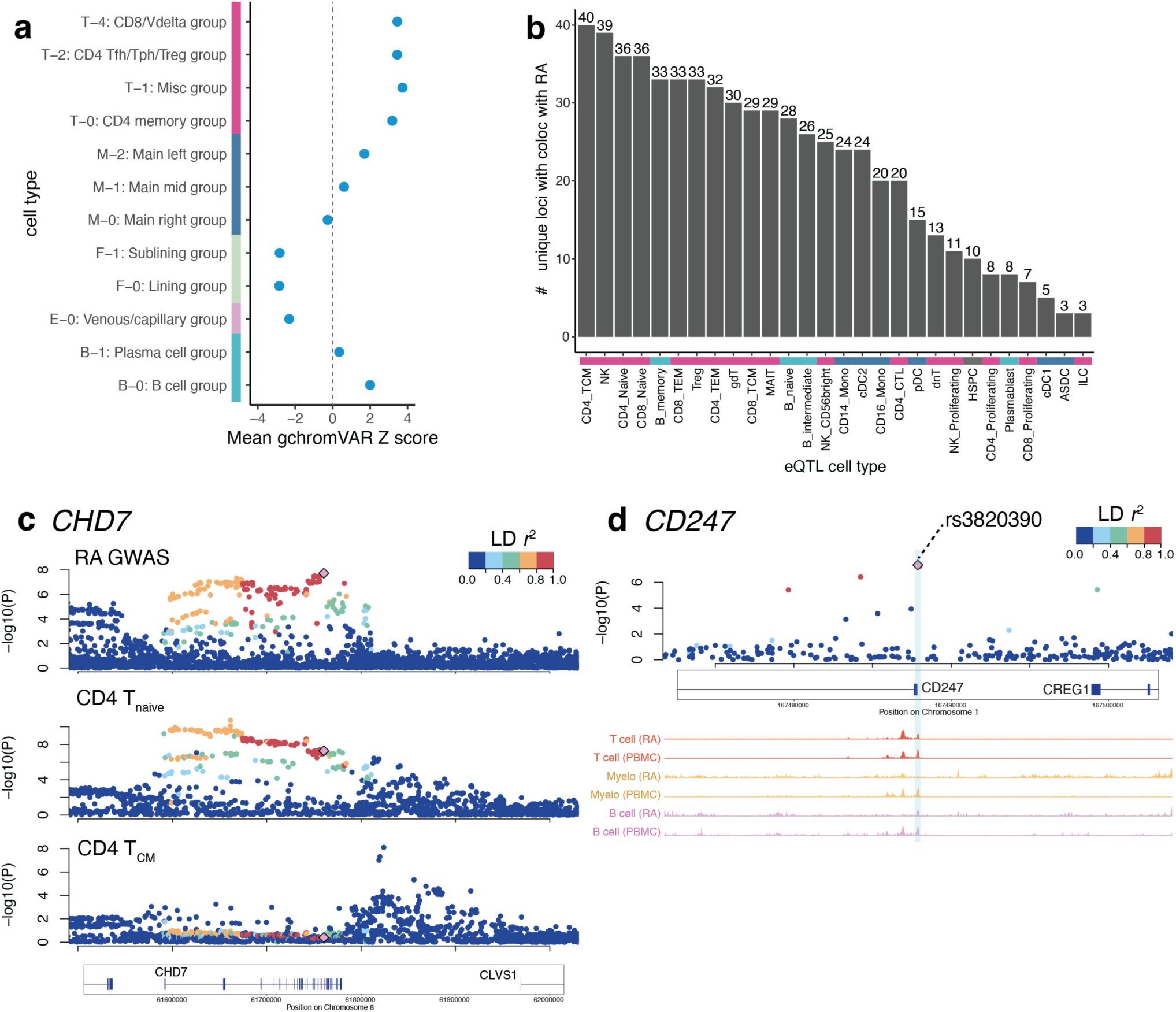
Functional fine-mapping of RA loci. **a.** g-chromVAR enrichment Z score of fine-mapped RA variants within cell-type-specific accessible chromatin regions. The bars next to the labels in the y-axis correspond to broad cell type categories. **b**. The number of unique loci that colocalized with the genome-wide significant loci in our RA GWAS in each single-cell eQTL cell type. The bars above the labels in the x-axis correspond to broad cell type categories. **c**. Novel *CHD7* locus colocalizing with single-cell CD4 naïve T cells eQTL. Regional plots for RA GWAS (top), CD4 naïve T cells (middle) and CD4 central memory T cells (bottom) are shown, with each SNP colored based on the LD from the lead variant in the GWAS as shown in the color scale. **d**. Novel *CD247* locus with a putative causal variant (rs3820390) at the SCENT promoter of the gene. Below the regional plot with SNPs colored based on the LD from the lead variant (color scale), cell-type-specific ATAC-seq accessibility tracks are shown including the functional promoter harboring the lead variant within a sky-blue band.

Second, to identify effector genes, we integrated single-cell expression quantitative trait loci (eQTL) with GWAS signals. We used eQTL maps of 28 immune cell types from peripheral blood in 1,925 individuals (TenK10K Phase 1), the largest single-cell atlas to date^42^. We identified 105 unique loci (74% of non-coding loci) with significant GWAS–eQTL colocalization^43^ (posterior probability (PP) of colocalization (H4)>0.7; **Supplementary Table 5**). Colocalizations were predominantly observed in T cell states, including central memory (TCM), naïve CD4⁺, and naïve CD8⁺ T cells (**Figure 4b**), although Tph cells were not represented in this peripheral blood dataset. 22 novel loci showed such colocalization, helping define cell-type-specific mechanisms. For example, *CHD7* emerged as a novel effector gene (**Figure 4c**) in naïve T cells (i.e., CD4+ and CD8+ naive T cells; PP(H4)>0.75). *CHD7* encoding a chromatin remodeler critical for multisystem development is a causative gene for CHARGE syndrome characterized by systemic congenital abnormalities including thymic hypoplasia or aplasia and T-cell immunodeficiency^44^. *ELMO1* was also identified as a new effector gene across T cell subsets (e.g., CD4+ and CD8+ T_CM_; PP(H4)>0.90) and has reported links to psoriasis^45^ and hypothyroidism^46^.

Third, we applied single-cell enhancer–gene mapping (SCENT)^47^ to functionally fine-map variants within 95% credible sets and their effector genes. Among novel seropositive loci, we identified a promoter variant (rs3820390; PIP=0.44; beta=-0.14, *P*=4.6×10⁻⁸) at *CD247* in T cells (**Figure 4d**). CD247 encodes CD3ζ chain essential for T cell receptor signaling and CD3 complex formation. *CD247* was previously reported for suggestive association for systemic lupus erythematosus. We also fine-mapped rs3768383 (PIP=0.13; P=6.0×10⁻¹¹) within an intronic enhancer at *ITPKB*, which regulates calcium signaling during T cell development. Supporting this, ITPKB eQTL colocalized with the GWAS signal in T_reg_s^42^ (PP(H4)=0.97). Outside T cells, we identified rs28504359 within a myeloid-specific enhancer targeting CD164, a sialomucin adhesion receptor involved in cell migration^48,49^. Overall, GWAS fine-mapped variants showed stronger enrichment in cell-type-specific regulatory elements than in previous GWAS, particularly in T cells (7.5X fold enrichment vs. 5.9X; **Supplementary Figure 5**).

We examined the chromatin accessibility and expression of the genes implicated in the RA GWAS. While T cells are well established in RA genetics^41,50,51^, other cell types are increasingly implicated in disease mechanisms. Clinically, CD20⁺ B cell depletion with rituximab or CD19×CD3 bispecific T cell engagers targeting both T and B cells can induce long-lasting remission^52,53^. Tissue-resident macrophages and fibroblast subpopulations are also expanded in specific RA subtypes, highlighting additional therapeutic targets^54^. To investigate these mechanisms, we used the joint single-cell profiling of chromatin and expression accessibility of synovial samples^39,54^. Genes at 42 loci (29%) were relatively highly expressed in T cells (Z score>0.5), while others showed higher expression in myeloid cells, dendritic cells, B cells, or fibroblasts (**Figure 5a**). For example, *FCRL3, CCR6, BLK*, and the novel locus *CHD7* were highly expressed in B cells, whereas *CH2B3, PILRA*, and novel loci *SPNS1* and *IL1RN* were enriched in myeloid lineages. In contrast, chromatin accessibility at RA loci was broadly shared across cell states (**Figure 5b**). This suggests that additional regulatory factors such as cell-type-specific transcription factor activity may drive the cell-state-specific expression of RA genes.

**Figure 5.**
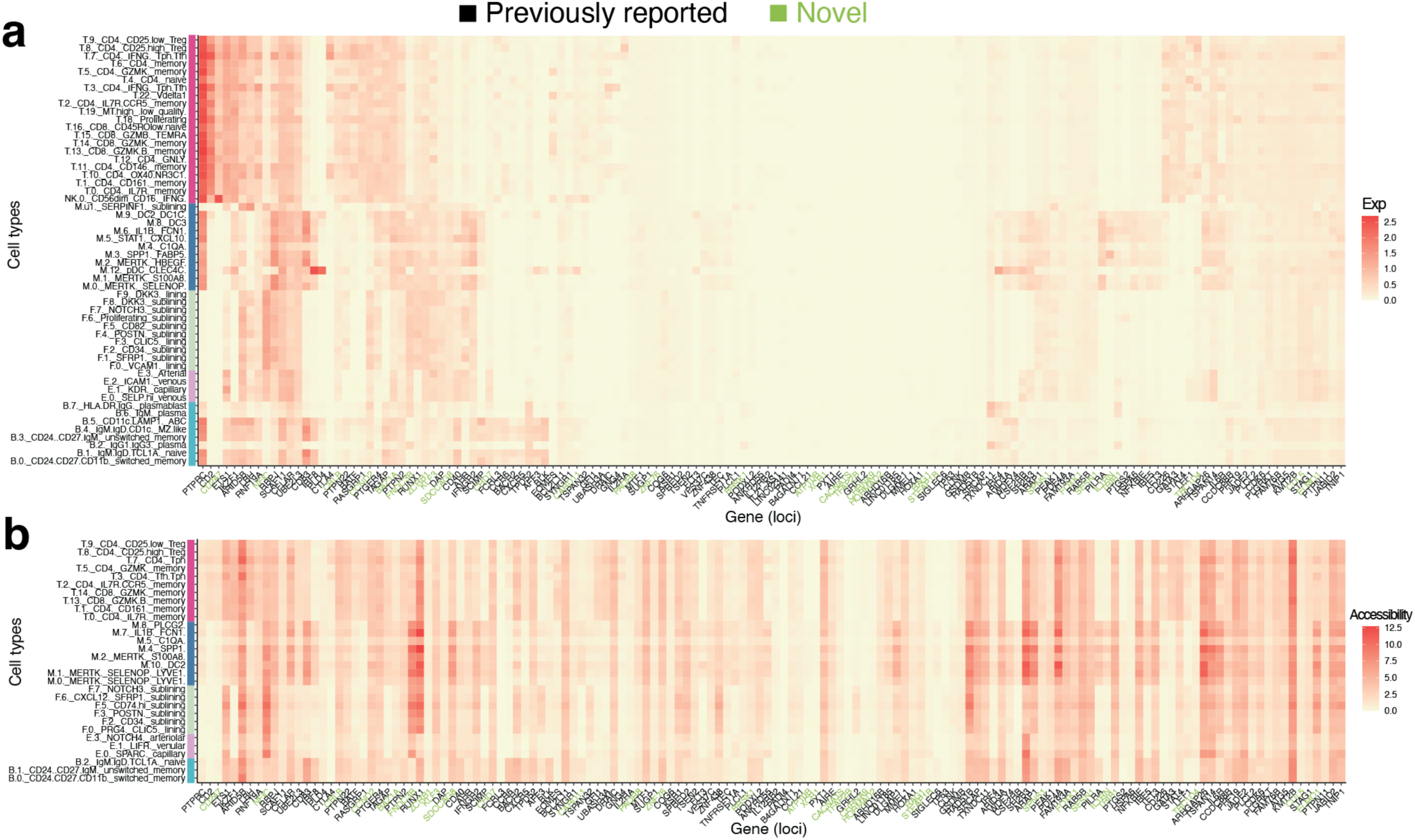
Cell-type characterization of RA loci. **a**. Cell-state specific expression of genes in RA GWAS loci using single-cell RNA-seq of RA synovial tissues. **b**. Cell-state specific chromatin accessibility (<10kb) of genes in RA GWAS loci using single-cell ATAC-seq of RA synovial tissues. Each of genome-wide significant loci was assigned to a most plausible functional gene (**Methods**) as in the x-axis. The loci are sorted by hierarchical clustering of gene expression (**a**) and we used the same order in **b**. Cell types that were assessed for expression and chromatin accessibility (y-axis) were colored by broad cell type classifications as colored in the bars.

To determine the broad phenotypic consequences of RA alleles, we investigated the association of the lead variants from our genome-wide significant loci across 1,263 health outcomes in the MVP (n_trait_=1,263). 48 variants showed genome-wide significant associations with 113 traits, including 31 immune-mediated conditions (**Supplementary Figure 6**; **Supplementary Table 6**). *SH2B3*, *PTPN22* and *PTPN11* loci were the most pleiotropic loci, associated with 84, 52, and 35 traits, respectively. Many associations unsurprisingly reflected shared autoimmune genetic risk^55^. The greatest overlap occurred with autoimmune thyroid diseases, including Graves’ disease and Hashimoto thyroiditis^56^, which were associated with 18 loci. Autoimmune thyroid disease and RA often co-occur in patients^57^. Some loci implicated other immunological diseases, such as psoriasis, for examples *PDE4A* and *UBE2L3*. The role of the immune system in cancer is now well-understood, and we observed associations with skin cancer, for example at *CTLA4* and *IL2RA.* Pleiotropic associations were also observed with ischemic heart disease and atherosclerosis, particularly at *IL6R*, *SH2B3*, and the novel locus *PTPN11*, highlighting shared inflammatory mechanisms in cardiovascular disease^58^.

The MHC, which includes the human leukocyte antigen (HLA) genes, contributes more heritability to RA than the rest of the genome^59^. Previous studies in EUR identified a strong independent association at the amino acid positions in the HLA-DRβ1 gene at positions 11 or 13, 71 and 74 and HLA-B at position 9^6^. Studies in EAS reported similar amino acid signals but heterogeneous effect sizes due to frequency differences^60^. Leveraging the larger AFR individuals in MVP, we performed HLA association tests and fine-mapping for seropositive RA. In AFR, the strongest association was observed at HLA-DRB1 position 11 (in complete LD with position 13; P=3.67×10^−33^), followed by a new position 85 (P=4.62×10^−3^; **Supplementary Figure 7a**). At HLA-B, position 9 was significantly associated (P = 1.27×10^−8^). In contrast in EUR samples in MVP, the strongest signal was observed at HLA-DRB1 position 11 (P < 10^−300^); the second association at 71 (P=4.81×10^−18^) and the third signal at 74 (P=4.77×10^−9^) were observed after conditioning, consistent with previous EUR studies. We considered the possibility that AFR and EUR have different secondary signals within the HLA. We replicated the AFR findings in three independent cohorts (CLEAR^61^, GENRA^62^ and Penn Medicine BioBank). Nominally significant haplotype effects (*P*<0.05) defined by AFR positions 11, 13, and 85 were consistent and directionally concordant (Peason’s correlation *r*=0.93 with *P*=8.0×10^−3^ in CLEAR and GENRA; *r*=0.93 with *P*=6.8×10^−3^ in Penn Medicine BioBank; **Figure 6a** and **6b**, see **Methods**). Some risk haplotypes are more frequent in AFR than in EUR, such as 11:D_13:F_85:V and 11:V_13:F_85:V (**Supplementary Figure 7b**).

**Figure 6.**
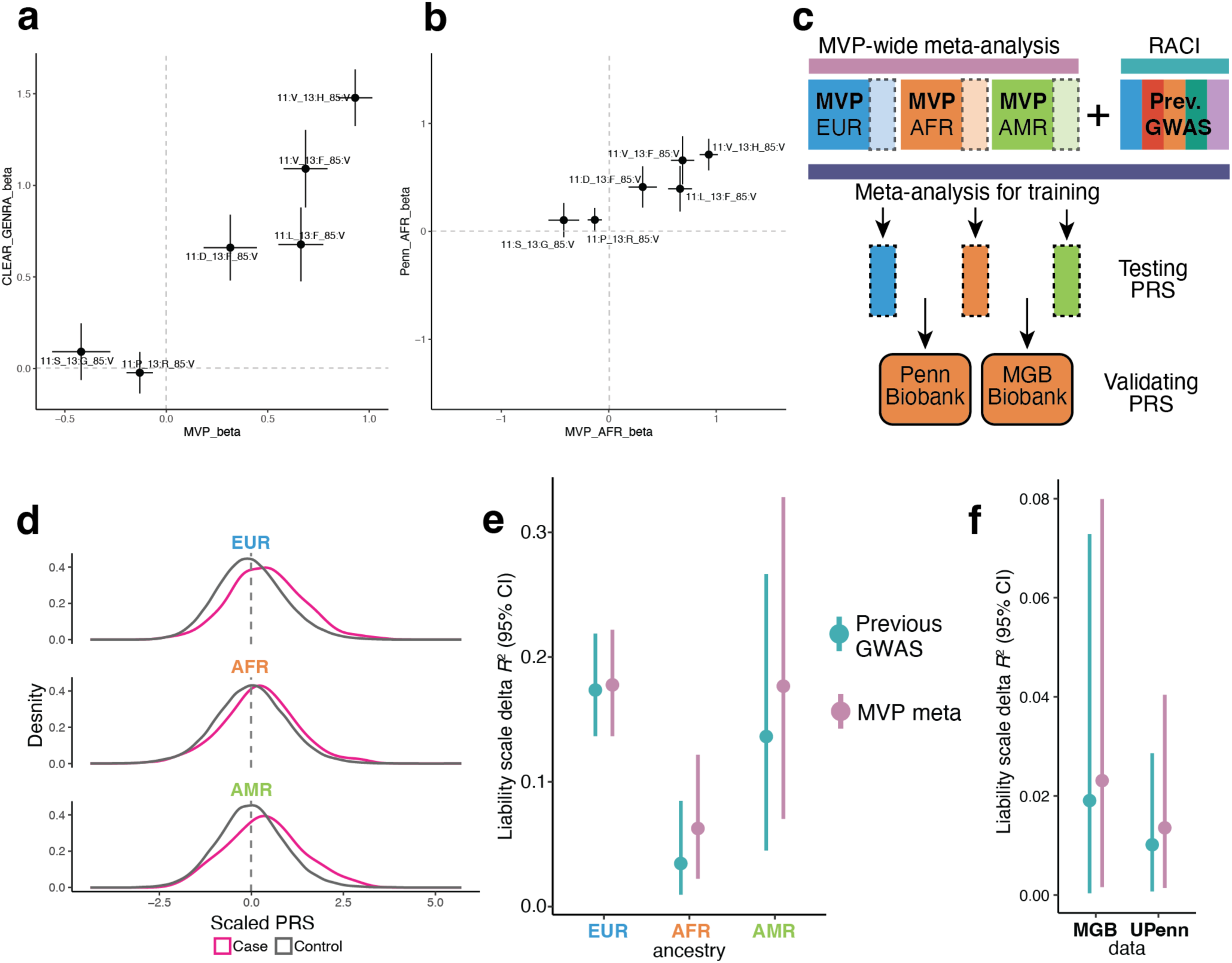
HLA and polygenic risk of RA. **a** and **b.** Replication of DRB1 HLA haplotype analyses using CLEAR and GENRA cohorts (**a**) and Penn Medicine BioBank (**b**). **c**. Our strategy to construct and validate RA PRS. **d**. Case and control distribution of PRS values in a testing dataset in MVP. **e**. Explained Nagerkerke’s *R^2^* in a liability scale in the testing dataset in MVP. **f**. Explained delta Nagerkerke’s *R^2^*in a liability scale in the validation datasets of MGB and Penn Biobanks.

Finally, we evaluated the performance of polygenic risk scores (PRSs) derived from our GWAS. We conducted an additional GWAS using 70% of MVP individuals followed by meta-analyses with RACI and trained the PRS weights using the remaining 30% (**Figure 6c**). In the held-out dataset, the PRS explained 17.8%, 6.3%, and 17.7% of phenotypic variance in a liability scale measured by Nagerkerke’s pseudo *R^2^*(**Methods**) in EUR, AFR, and AMR, respectively (**Figure 6d** and **6e**). While performance in EUR remained unchanged from previous GWAS Ishigaki et al.^3^ (17.8% vs 17.4%), we observed improvements in AFR (6.3% vs 3.3%) and AMR (17.7% vs 13.6%), although the confidence intervals overlapped. We further validated PRS performance in external AFR datasets, Penn Medicine BioBank^63^ and Mass General Brigham (MGB) Biobank^64^. Our PRS explained 1.4% of phenotypic variance in AFR in Penn Medicine BioBank vs 1.0% using the previous RA GWAS PRS and 2.3% vs 1.9% in MGB Biobank (**Figure 6f**) again with overlapping confidence intervals. Overall, inclusion of AFR and AMR samples along with the improved PRS performance in underrepresented populations, with no differences in the already well-represented EUR population.

## Discussion

We presented the largest cross-ancestry meta-analysis GWAS of RA by leveraging the size and diversity of MVP with the largest AFR and AMR populations to date. We identified 31 novel loci powered by increased sample size and population representation, revealing new biological insights into RA pathogenesis. Statistical fine-mapping reduced credible sets aided by ancestry-specific LD differences, and implicated non-coding regulatory putative causal variants functioning in immune cell types.

RA is a challenging phenotype to study in the context of biobanks, given its relatively low prevalence, and the low accuracy of RA diagnostic codes in classifying RA with EHR-based data^65^. The improved accuracy of the RA phenotype with the application of machine learning pipelines designed for use with EHR data improved the power to detect associations and can be easily transported for use in other EHR linked biobanks.

Combining more refined sets of the RA GWAS variants with the functional epigenomic and transcriptomic resources at the single-cell resolution, we elucidated granular cell states, such as naïve CD4+ T cells, that RA alleles function to alter gene regulation. We nominated novel target genes of the newly identified and known RA alleles using approaches of colocalization with eQTL and cell-type-specific enhancer-gene maps at the single-cell resolution (e.g., *CHD7* and *CD247*) that could potentially serve as candidates for therapeutic target. At the HLA locus, AFR-specific analyses identified a potential novel amino acid position in HLA-DRB1 associated with RA risk. Finally, our inclusive GWAS improved PRS performance in underrepresented populations. We publicly released these summary statistics to further promote novel hypothesis generation in the RA biology.

## Data Availability

GWAS summary statistics for MVP will be available through dbGaP (https://www.ncbi.nlm.nih.gov/gap/) at study accession phs001672 (in progress). The meta-analyses data of MVP with RACI can be found on VA CIPHER (https://phenomics.va.ornl.gov/web/cipher/partner/mvp).

## Code Availability

The computational scripts related to this manuscript are available on GitHub (https://github.com/Sakaue-Lab/MVP-RA-GWAS).

## Methods

### Study participants of MVP

The VA Million Veteran Program (MVP) is a national United States cohort initiating enrollment in 2011 to determine the contributions of genetics, lifestyle, and military exposures to health and disease among US Veterans^15,18^. Blood specimens from participants were collected for genotyping. The biorepository was linked with the VA electronic health record (EHR), which includes diagnosis codes (International Classification of Diseases ninth revision [ICD-9] and tenth revision [ICD-10]), laboratory measures, and detailed survey questionnaires collected at the time of enrollment for all Veterans and followed in the healthcare system until September 2020.

### Genotyping and genome-wide imputation in the MVP

Genotyping and imputation methods for the MVP were described previously^66^. In brief, the single nucleotide polymorphism (SNP) data in the MVP cohort were generated using a custom ThermoFisher Axiom MVP 1.0 genotyping platform. Variants were then imputed using the QCed MVP genotypes and TOPMed imputation reference panel^67^. Following imputation, variant level quality control (QC) was performed, and genetic variants with (1) imputation quality <0.3, (2) minor allele count (MAC) <20, (3) call rate <97.5% for common variants (minor allele frequency [MAF] >1%), and (4) call rate <99% for rare variants (MAF<1%) were excluded.

### Phenotyping of RA in the MVP

The current standard for phenotyping in most biobanks with linked EHR is to classify based on billing codes, e.g., ICD10. However, for some conditions, such as RA, billing codes alone can only achieve modest positive predictive value^24–26,65^. To address this challenge, we applied an unsupervised machine learning approach, knowledge-driven online multimodal automated phenotyping (KOMAP)^27^, combining structured EHR data, (e.g., ICD codes, medication prescriptions) with natural language processing EHR to improve the accuracy of phenotyping and improve the power for an association study^27^. The performance of the KOMAP derived RA phenotype was validated with manual chart review. The primary definition of RA in the MVP RA GWAS was reported using RA classified with KOMAP. An RA GWAS was performed in MVP using the standard definition of ≥2 RA ICD codes to compare with the results of the GWAS results using KOMAP. Patients with exactly 1 RA ICD code were removed from the analysis, consistent with the practice in previous studies.

### Population assignment in the MVP

We used previously estimated population group membership as described in detail in ref^18^. In brief, we obtained a reference dataset from 1000G and used the smartpca module in the EIGENSOFT package to project the PC loadings from a group of unrelated individuals in the reference dataset. We merged this dataset with the MVP dataset and ran smartpca to project the PCA loadings from the reference dataset. We trained a random forest classifier using continental ancestry meta-data based on the top 10 principal components from the reference training data to define genetically inferred ancestry. We then applied this random forest to the predicted MVP PCA data and assigned populations to individuals with a probability greater than 50%. Those with a probability less than 50% for any particular population group were excluded from the study. Figure S11 shows the PCA projection of MVP participants on the 1000G reference panel. Individuals were assigned to populations groups defined as genetically similar to African (AFR), Admixed American (AMR), East Asian (EAS), European (EUR), and South Asian (SAS). Only AFR, AMR, and EUR had enough cases to be included (case count > 50) in these analyses. Lastly, we selected PC1-5 to adjust our analysis.

### GWAS in the MVP

Within each population group (AFR, AMR and EUR) and in each phenotypic category, we performed a GWAS with the linear mixed model method implemented in REGENIE^68^, a two-step whole-genome regression framework that accounts for sample relatedness and unobserved population structure. Only variants with an imputation quality *Rsq* >0.5 and MAF >0.1% within the relevant population group were included in the GWAS. Analyses were adjusted for age, sex, and five population-specific genetic PCs.

### Meta-analyses in the MVP and with previous GWAS cohorts (RACI)

We used METAL^69^ for performing the fixed-effect, inverse-variance weighted meta-analyses across three ancestries within the MVP and also performing the meta-analyses between the MVP and our previously conducted GWAS from RACI, including 37 cohorts consisting of 35,871 RA cases and 240,149 control individuals of EUR, EAS, AFR, SAS and ARB ancestry^3^. We also conducted secondary GWAS only using EUR samples and seropositive RA samples and controls.

### Estimation of heritability and bias in GWAS results

We used S-LDSC using the baselineLD model (v2.1) and LDSC software (v1.0.0)^23^ to estimate heritability and confounding bias in EUR GWAS (MVP only and meta-analysis with RACI) in which we have sufficient statistical power to perform this analysis, since S-LDSC assumes that GWAS has samples from a single ancestral background and a sufficient sample size. We estimated that the prevalence of RA was 0.5% as conventionally used^3^ to calculate liability-scale heritability from observed-scale heritability.

### Determination of significant loci and independently associated SNPs

Genome-wide significant threshold was set at P < 5×10^−8^ as conventionally used in previous GWAS studies. We defined a locus as a genomic region within ±1 megabase (Mb) from the lead variant, and we considered a locus as novel when it did not include any variants previously reported for RA, with the exception of MHC region in which we more closely investigated the LD and association patterns to define independent loci given their extended LD structure. We defined reported variants as significant variants (*P* < 5 × 10^−8^) reported in the GWAS Catalog (https://www.ebi.ac.uk/gwas) accessed on May 15, 2025.

### Statistical fine-mapping analyses

We conducted fine-mapping analysis using approximate Bayesian factor (ABF) and constructed 95% credible set for each significant locus that we defined as described above, excluding the MHC region. As inputs, we used the summary statistics of each ancestral composition and seropositive status in which we observed genome-wide significance. We calculated ABF of each variant according to ref^36,43,70^:

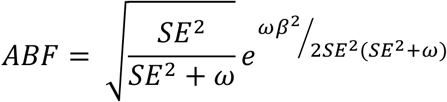

where 𝛽 and SE are the variant’s effect size and standard error, respectively; 𝜔 denotes the prior variance in allelic effects (we empirically set this value to be 0.04) from the ref^36^. For each locus, we calculated the posterior inclusion probability (PIP) of variant *k* according to:

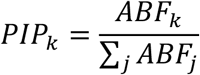

where *j* denotes all of the variants included in the locus. We sorted all variants in the order of decreasing PIP and constructed 95% credible set (CS) including variants from the top PIP until the cumulative PIP reached 0.95.

### Colocalization with single-cell eQTL data

To investigate the sharing of causal variants between RA GWAS loci and cell-type-specific eQTL in peripheral blood immune cells, we downloaded summary statistics of single-cell eQTL from TenK10K Phase 1 (https://zenodo.org/records/17474113). We used the full summary statistics for common variant *cis* eQTL mapping for the 28 immune cell types in PBMC. For each RA loci, we extracted summary statistics with a +/- 500kb window centered around the lead variant, and analyzed any genes that overlapped with any of these regions and had more than 200 variants intersected with the corresponding GWAS locus. We used coloc software (version 5.2.3) to perform fine-mapping and colocalization analyses using ABF between RA GWAS and eQTL from each cell type. We defined significant colocalization when we observed posterior probability (PP) for hypothesis 4 (H4; a shared causal variant) > 0.7.

### Functional fine-mapping using SCENT

To jointly fine-map causal variants and genes, we used single-cell enhancer-gene mapping (SCENT)^47^ which uses single-cell multiome RNA/ATAC data to connect cell-type-specific functional enhancers to their target genes. We used previously constructed T, B, and myeloid cell-type specific enhancer-gene maps across 7 multiome datasets^39,71–74^. We lifted our meta-analysis fine-mapped variants within 95% CS over to hg38 and identified putative statistically fine-mapped causal variants within the SCENT enhancers with jointly identifying the target genes in each cell type. The overall enrichment of RA variants within cell-type-specific enhancers were calculated as ref.^47^:

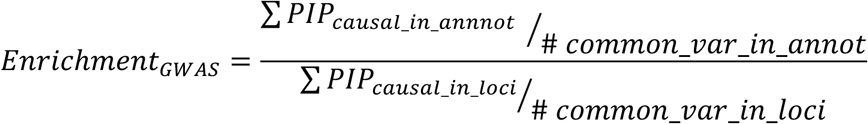

For our RA GWASs (either our MVP meta-analysis or previous Ishigaki et al.), we divided the sum of PIPs from fine-mapped putatively causal variants with PIP>0.2 within an annotation (across all genome-wide loci) normalized by the number of common variants within an annotation by the sum of PIPs from all fine-mapped putatively causal variants normalized by the number of all common variants within all significant loci. To calculate common variants within annotation or within locus, we again used 1000 Genomes Project variants with minor allele frequency > 1% in European population.

### Single-cell expression and chromatin accessibility

We used previously published single-cell gene expression^54^ and chromatin accessibility^39^ catalogs of synovial tissues from RA and osteoarthritis patients. We combined evidence for the target gene of each of our 152 genome-wide significant loci, including the closest genes, coding genes, eQTL colocalization and functional enhancer-gene maps. For these genes, we pseudobulked log-normalized gene expression per cell state annotated in the ref.^54^. The expression information on 144 genes was obtained in total. In addition, we scaled the gene expression by deriving Z scores to investigate the relative expression across cell states. From ATAC chromatin accessibility data from single-cell multiome (RNA/ATAC) profiling of synovial tissues^39^, we extracted 200 bp peaks within 10 kb of transcription start sites of the same genes we defined as above and created a pseudobulked accessibility profile across cell states annotated previously with both RNA and ATAC.

### g-chromVAR cell state enrichment

We lifted our GWAS fine-mapped variants in 95% CS over to GRCh38. We re-processed single-cell multiome (RNA/ATAC) profiling of synovial tissues and aggregated chromatin peak counts in 12 immune and non-immune populations with the number of cells > 500. We used gchromVAR software (v0.3.2) to investigate enrichment of fine-mapped SNPs within these accessible regions in a cell state with default parameters. We repeated the same analysis for 1000 times to derive the mean and standard error of enrichment Z scores in each cell type.

### HLA association analyses

We imputed the classical HLA alleles (up to two fields) and corresponding amino acid sequences of MVP samples with the QCed genotype on chromosome 6 and a haplotype reference panel consisted of 1KGP samples using a modified version of SNP2HLA^59,75^ which uses Minimac3^76^ instead of Beagle for scalablity. We applied post-imputation quality control to keep the imputed variants with minor allele frequency (MAF) > 1% and Rsq > 0.7 and removed individuals having any *HLA* gene for which the total number of two-field alleles does not sum up to exactly 2 (<1.95 or >2.05). Using imputed dosages from the HLA imputation, we performed a series of omnibus and conditional haplotype tests^6,59^ to identify independent amino acid residue positions that significantly influence RA risk. First by an omnibus test for an amino acid position which has 𝑀 possible amino acid residues, we assessed the significance of the improvement in fit for the full model which includes 𝑀 − 1 amino acid dosages as explanatory variables when compared to a reduced model without those amino acid dosages. We assessed the improvement in model fit by the delta deviance (sum of squares) using an F-test with 𝑀 − 1 degrees of freedom and derive the statistical significance of the improvement.

Full model: 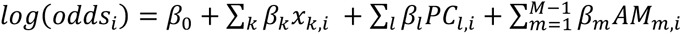

Reduced model: 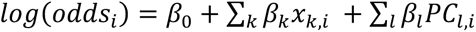
where 𝑚 is one amino acid residue at this position, 𝑀 is the total number of observed amino acid residues at this position, 𝐴𝑀_;,5_ and 𝛽_;_are the amino acid dosage of the residue 𝑚 in individual 𝑖 and the effect size for the residue 𝑚, respectively. In this analyses, we focused on seropositive RA as a phenotype given the heterogeneity of HLA risk between seropositive and seronegative RA.

Next we grouped all possible two-field alleles for this *HLA* gene into 𝑀 groups based on the most significant amino acid position and estimated the effect of each of the 𝑀 groups using a logistic regression model. We assessed the improvement in model fit over a reduced model without including those *M* groups by the delta deviance using an F-test with 𝑀 − 1 degrees of freedom and derive the statistical significance of the improvement.

Full model: 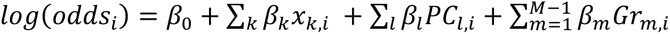

Reduced model: 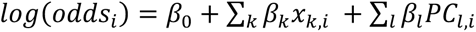
where 𝐺𝑟*_m,i_* is the sum of the dosage of two-field alleles from a group 𝑚, explained by the 𝑚’th amino acid residue.

Finally we sought to identify which amino acid position other than this significant position best improves the model over the model only including this significant position. Let 𝑥 be the most significant position in the primary analysis, which has 𝑋 possible amino acid residues. We sequentially test each amino acid position (𝑧) other than 𝑥, to ask whether haplotypes defined by the amino acid combination of positions 𝑥 and 𝑧 (𝑧 ≠ 𝑥) explain the disease risk more than those defined only by the position 𝑥. To do so, we re-categorize all two-field alleles at this *HLA* gene into 𝑍 groups, where 𝑍 is the total number of observed haplotypes defined by the amino acid positions 𝑥 and 𝑧. The value of 𝑍 must be at least 𝑋 if no new haplotypes are defined. We again assess the significance of the improvement in model fit of the Full model (covariation at positions 𝑥 and 𝑧) over the Reduced model (variation at position 𝑥 alone) by the delta deviance (sum of squares) using an F-test with (𝑍 − 𝑋) degrees of freedom.

Full model: 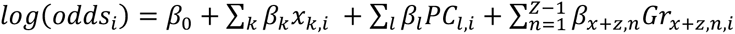

Reduced model: 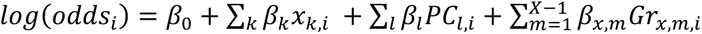

where 𝐺𝑟*_x+z,n,i_* is the sum of the dosages of two-field alleles in a group 𝑛 by a given combination of the amino acid residues at positions 𝑥 and 𝑧. Thus, we define the next most significant amino acid position which additionally and independently explains the disease risk from the position 𝑥. If the model improvement in this second round is statistically significant, we iterate the same analyses to identify amino acid position(s) other than the previously identified positions that best improve the model over the model including those previous positions, until we obtain no further significant improvement from any of the remaining positions. To visualize molecular positions of our intendent positions from conditional haplotype tests, we used UCSF chimera based on Protein Data Bank entries 3pdob (**Supplementary Figure 7a**).

In the replication cohorts of Penn Medicine Biobank (381 cases, 11,660 controls), GENRA (166 cases, 867 controls), and CLEAR (348 cases, 1,004 controls) cohorts, we directly constructed haplotypes from independent amino acid positions in the MVP and assessed their effects on seropositive RA risk using sex, age, and genotype PCs as covariates. The CLEAR registry enrolled AAs with RA in two phases. CLEAR 1 enrolled AAs of ≤2 years disease duration (from 2000 to 2006) and CLEAR 2 enrolled AAs with RA (not previously enrolled in CLEAR 1) of any disease duration (2006–2011). Participants were enrolled at the following academic sites: University of Alabama at Birmingham (Coordinating Center); Grady Hospital/Emory University, Atlanta, GA; University of North Carolina, Chapel Hill, NC; Washington University, St. Louis, MO; and Medical University of South Carolina, Charleston, SC. CLEAR controls were AAs without rheumatic disease who were age-, sex- and geographic location-matched (as a group) to the CLEAR RA patients. The GENRA included people of African ancestry living in the UK (Black African, Black Caribbean or Black British ethnicity). The GENetics of RA in individuals of African ancestry (GENRA) study which recruited patients with RA was approved by the National Research Ethics Service Committee London—Dulwich (reference: 11/LO/1244). Controls for the GENRA study were from the South London Ethnicity and Stroke Study (SLESS); this was approved by the Wandsworth Local Research Ethics Committee (reference: 05/Q0803/324). All participants provided consent.

### Phenome-wide association studies

We looked up previously generated GWAS summary statistics across 1,263 binary disease traits in the MVP for our genome-wide significant lead variants. Since the PheWAS results available for look-up were based on the MVP genotype based on imputation using 1KGP reference panel, not all of the genome-wide lead variants were able to be assessed. Therefore 147 loci among 152 loci were analyzed in total. In preparing the **Supplementary Figure 6**, we calculated the Z scores from beta and standard error in GWAS and assigned the sign of the Z score as positive if the effect direction is consistent with RA and negative if it is opposite.

### Polygenic risk score

We used the pruning and thresholding method to calculate the PRS in this study. The study was designed to ensure that the samples used in constructing PRS were completely independent from the samples used in the validation; we performed an additional GWAS using 70% of MVP individuals followed by meta-analyses with RACI. With the beta from this GWAS as PRS weights, we used the remaining 30% of MVP samples to choose the PRS parameters (i.e., P value thresholds and r2 thresholds) that maximized the explained RA variance. For LD-pruning, we used haplotype information in a subset of randomly selected EUR samples from the MVP (n=1,000). We used ten different P value thresholds: 0.1, 0.03, 0.01, 0.003, 0.001, 3.0 × 10^−4^, 1.0 × 10^−4^, 3.0 × 10^−5^, 1.0 × 10^−5^ and 5.0 × 10^−8^. We assessed each of PRS weights by comparing Nagelkerke’s *R^2^*with including or excluding the PRS in a logistic regression model with the same covariates as used in GWAS. After choosing the parameters for PRS, we compared the explained RA variance by Nagelkerke’s *R^2^* between our PRS and that from the previous GWAS in Ishigaki et al^3^. Note that Ishigaki et al. also published a functionally informed PRS using epigenomic annotations, however, our PRS was purely derived from the pruning and thresholding to allow for a fair comparison. We converted this observed scale delta *R^2^* into liability scale with using a population prevalence of RA being 0.5% as conventionally used.

We tested the performance of our PRS using the following independent AFR datasets: Penn Medicine BioBank (381 cases, 11,660 controls) and MGB biobank (91 cases, 1,442 controls). Note that the AFR ancestry cohorts, GENRA and CLEAR that we used in the HLA analyses, were already included in the previous Ishigaki et al. RA GWAS and were therefore not included in the PRS validation to maintain the independence among cohorts. In each AFR cohort, we again compared the explained RA variance between our PRS and that from the previous RA GWAS by Ishigaki et al. using Nagelkerke’s *R^2^*. We converted the observed scale delta *R^2^* into liability scale with using a population prevalence of RA being 0.5% as conventionally used.

## Acknowledgments

This study was conducted with data from the VA Million Veterans Program which is supported by the Office of Research and Development, Veterans Health Administration. This study was supported by MVP001 as well as award # BX004821 and MVP000. Thank you to the MVP participants for their service and their continued contributions to our nation through participation in MVP. The opinions expressed in this article are those of the authors and do not necessarily represent those of the Department of Veterans Affairs or the United States government. GENRA was funded by Arthritis Research UK (Grant Reference 19739 to ICS). We appreciate the RACI for providing us with the summary statistics from the international RA GWAS.

## Author Contributions

S.S., T.C., S.R., and K.P.L. conceived the work and wrote the manuscript with inputs from all the other authors. D.P., Z.R., Z.L, J.C., A.B., Y.H., L.C., P.M., S.H., K.I, C.M., V.T., R.S., M.M, S.M., V.P., G.M., J.S.H., T.R., V.L., Y.O., I.S., S.L.B, J.B. contributed to data acquisition and curation. S.S., D.Y., H.Z., D.P., C.H., T.C., K.P.L. contributed to RA phenotyping, the MVP GWAS, meta-analyses, and downstream statistical and functional analyses. Z.R. and A.V. performed replication analyses at Penn Medicine Biobank. P.W.W., J.M.G., C.H., A.V., K.C., J.E.H., T.C., S.R., K.P.L had supervisory roles.

## Competing Financial Interests

S.S. declares no competing interests. S.R. is a founder for Mestag, Inc, a scientific advisor for Jannsen, and Pfizer, and serves as a consultant for Sanofi, Abbvie, Nimbus and Third Rock Ventures. K.P.L. served as a consultant for BMS, Merck, and UCB outside of the scope of this work.

## Notes

### Author Declarations

IRB of the Department of Veteran Affairs (VA) gave ethical approval for this work (Million Veterans Program MVP001, CA cIRB 10-02 and 16-06). IRB of Brigham and Women's Hospital gave ethical approval for this work, #2020P002053.

## References

1. Gravallese, E.M., and Firestein, G.S. (2023). Rheumatoid Arthritis - Common Origins, Divergent Mechanisms. N. Engl. J. Med. 388, 529–542. 10.1056/nejmra2103726.

2. Macgregor, A.J., Snieder, H., Rigby, A.S., Koskenvuo, M., Kaprio, J., Aho, K., and Silman, A.J. (2000). CHARACTERIZING THE QUANTITATIVE GENETIC CONTRIBUTION TO RHEUMATOID ARTHRITIS USING DATA FROM TWINS. Arthritis Rheum. 43, 30–37. 10.1002/1529-0131(200001)43:1<30::AID-ANR5>3.0.CO;2-B.

3. Ishigaki, K., Sakaue, S., Terao, C., Luo, Y., Sonehara, K., Yamaguchi, K., Amariuta, T., Too, C.L., Laufer, V.A., Scott, I.C., et al. (2022). Multi-ancestry genome-wide association analyses identify novel genetic mechanisms in rheumatoid arthritis. Nat. Genet. 54, 1640–1651. 10.1038/S41588-022-01213-W.

4. Amariuta, T., Luo, Y., Gazal, S., Davenport, E.E., van de Geijn, B., Ishigaki, K., Westra, H.J., Teslovich, N., Okada, Y., Yamamoto, K., et al. (2019). IMPACT: Genomic Annotation of Cell-State-Specific Regulatory Elements Inferred from the Epigenome of Bound Transcription Factors. Am. J. Hum. Genet. 104, 879–895. 10.1016/j.ajhg.2019.03.012.

5. Gregersen, P.K., Silver, J., and Winchester, R.J. (1987). The shared epitope hypothesis. An approach to understanding the molecular genetics of susceptibility to rheumatoid arthritis. Arthritis Rheum. 30, 1205–1213. 10.1002/art.1780301102.

6. Raychaudhuri, S., Sandor, C., Stahl, E.A., Freudenberg, J., Lee, H.S., Jia, X., Alfredsson, L., Padyukov, L., Klareskog, L., Worthington, J., et al. (2012). Five amino acids in three HLA proteins explain most of the association between MHC and seropositive rheumatoid arthritis. Nat. Genet. 44, 291–296. 10.1038/ng.1076.

7. Okada, Y., Wu, D., Trynka, G., Raj, T., Terao, C., Ikari, K., Kochi, Y., Ohmura, K., Suzuki, A., Yoshida, S., et al. (2013). Genetics of rheumatoid arthritis contributes to biology and drug discovery. Nature 506, 376. 10.1038/nature12873.

8. Welter, D., MacArthur, J., Morales, J., Burdett, T., Hall, P., Junkins, H., Klemm, A., Flicek, P., Manolio, T., Hindorff, L., et al. (2014). The NHGRI GWAS Catalog, a curated resource of SNP-trait associations. Nucleic Acids Res. 42, D1001–6. 10.1093/nar/gkt1229.

9. Buniello, A., Macarthur, J.A.L., Cerezo, M., Harris, L.W., Hayhurst, J., Malangone, C., McMahon, A., Morales, J., Mountjoy, E., Sollis, E., et al. (2019). The NHGRI-EBI GWAS Catalog of published genome-wide association studies, targeted arrays and summary statistics 2019. Nucleic Acids Res. 47, D1005–D1012. 10.1093/NAR/GKY1120.

10. Greenberg, J.D., Spruill, T.M., Shan, Y., Reed, G., Kremer, J.M., Potter, J., Yazici, Y., Ogedegbe, G., and Harrold, L.R. (2013). Racial and Ethnic Disparities in Disease Activity in Patients with Rheumatoid Arthritis. Am. J. Med. 126, 1089–1098. 10.1016/j.amjmed.2013.09.002.

11. Suarez-Almazor, M.E., Berrios-Rivera, J.P., Cox, V., Janssen, N.M., Marcus, D.M., and Sessoms, S. (2007). Initiation of disease-modifying antirheumatic drug therapy in minority and disadvantaged patients with rheumatoid arthritis. J. Rheumatol. 34.

12. Yip, K., and Navarro-Millan, I. (2021). Racial, Ethnic, and Healthcare Disparities in Rheumatoid Arthritis. Curr. Opin. Rheumatol. 33, 117. 10.1097/BOR.0000000000000782.

13. Chen, M.H., Raffield, L.M., Mousas, A., Sakaue, S., Huffman, J.E., Moscati, A., Trivedi, B., Jiang, T., Akbari, P., Vuckovic, D., et al. (2020). Trans-ethnic and Ancestry-Specific Blood-Cell Genetics in 746,667 Individuals from 5 Global Populations. Cell 182, 1198–1213.e14. 10.1016/j.cell.2020.06.045.

14. Martin, A.R., Kanai, M., Kamatani, Y., Okada, Y., Neale, B.M., and Daly, M.J. (2019). Clinical use of current polygenic risk scores may exacerbate health disparities. Nat. Genet. 51, 584–591. 10.1038/s41588-019-0379-x.

15. Gaziano, J.M., Concato, J., Brophy, M., Fiore, L., Pyarajan, S., Breeling, J., Whitbourne, S., Deen, J., Shannon, C., Humphries, D., et al. (2016). Million Veteran Program: A mega-biobank to study genetic influences on health and disease. J. Clin. Epidemiol. 70, 214–223. 10.1016/j.jclinepi.2015.09.016.

16. Bycroft, C., Freeman, C., Petkova, D., Band, G., Elliott, L.T., Sharp, K., Motyer, A., Vukcevic, D., Delaneau, O., O’Connell, J., et al. (2018). The UK Biobank resource with deep phenotyping and genomic data. Nature 562, 203–209. 10.1038/s41586-018-0579-z.

17. Bick, A.G., Metcalf, G.A., Mayo, K.R., Lichtenstein, L., Rura, S., Carroll, R.J., Musick, A., Linder, J.E., Jordan, I.K., Nagar, S.D., et al. (2024). Genomic data in the All of Us Research Program. Nature 2024 627:8003 627, 340–346. 10.1038/s41586-023-06957-x.

18. Verma, A., Huffman, J.E., Rodriguez, A., Conery, M., Liu, M., Ho, Y.L., Kim, Y., Heise, D.A., Guare, L., Panickan, V.A., et al. (2024). Diversity and scale: Genetic architecture of 2068 traits in the VA Million Veteran Program. Science (1979). *385*. 10.1126/science.adj1182.

19. Whitbourne, S.B., Williams, A.R., Brewer, J. V., Deen, J.E., Lord, E.M., Murphy, S.A., Li, Y., Ho, Y.-L., Pyatt, M., Wolfrum, S., et al. (2025). Overview of Recruitment Strategies to Reach the Million Milestone and Characterization of the Million Veteran Program Cohort. 10.21203/rs.3.rs-8272722/v1.

20. Hunter, T.M., Boytsov, N.N., Zhang, X., Schroeder, K., Michaud, K., and Araujo, A.B. (2017). Prevalence of rheumatoid arthritis in the United States adult population in healthcare claims databases, 2004-2014. Rheumatol. Int. 37, 1551–1557. 10.1007/s00296-017-3726-1.

21. Myasoedova, E., Crowson, C.S., Kremers, H.M., Therneau, T.M., and Gabriel, S.E. (2010). Is the incidence of rheumatoid arthritis rising?: results from Olmsted County, Minnesota, 1955-2007. Arthritis Rheum. 62, 1576–1582. 10.1002/art.27425.

22. Zheng, G., Freidlin, B., and Gastwirth, J.L. (2006). Robust Genomic Control for Association Studies. The American Journal of Human Genetics 78, 350–356. 10.1086/500054.

23. Bulik-Sullivan, B., Loh, P.R., Finucane, H.K., Ripke, S., Yang, J., Patterson, N., Daly, M.J., Price, A.L., Neale, B.M., Corvin, A., et al. (2015). LD score regression distinguishes confounding from polygenicity in genome-wide association studies. Nat. Genet. 47, 291–295. 10.1038/ng.3211.

24. Wei, W.Q., Teixeira, P.L., Mo, H., Cronin, R.M., Warner, J.L., and Denny, J.C. (2015). Combining billing codes, clinical notes, and medications from electronic health records provides superior phenotyping performance. J. Am. Med. Inform. Assoc. 23, e20. 10.1093/jamia/ocv130.

25. Grams, M.E., Waikar, S.S., MacMahon, B., Whelton, S., Ballew, S.H., and Coresh, J. (2014). Performance and limitations of administrative data in the identification of AKI. Clinical Journal of the American Society of Nephrology 9, 682–689. 10.2215/CJN.07650713.

26. Cai, T., Zhang, Y., Ho, Y.L., Link, N., Sun, J., Huang, J., Cai, T.A., Damrauer, S., Ahuja, Y., Honerlaw, J., et al. (2018). Association of Interleukin 6 Receptor Variant With Cardiovascular Disease Effects of Interleukin 6 Receptor Blocking Therapy: A Phenome-Wide Association Study. JAMA Cardiol. 3, 849–857. 10.1001/jamacardio.2018.2287.

27. Xiong, X., Sweet, S.M., Liu, M., Hong, C., Bonzel, C.-L., Ayakulangara Panickan, V., Zhou, D., Wang, L., Costa, L., Ho, Y.-L., et al. (2025). Knowledge-Driven Online Multimodal Automated Phenotyping System. medRxiv. 10.1101/2023.09.29.23296239.

28. Lek, M., Karczewski, K.J., Minikel, E. V., Samocha, K.E., Banks, E., Fennell, T., O’Donnell-Luria, A.H., Ware, J.S., Hill, A.J., Cummings, B.B., et al. (2016). Analysis of protein-coding genetic variation in 60,706 humans. Nature 536, 285–291. 10.1038/nature19057.

29. Karczewski, K.J., Francioli, L.C., Tiao, G., Cummings, B.B., Alföldi, J., Wang, Q., Collins, R.L., Laricchia, K.M., Ganna, A., Birnbaum, D.P., et al. (2020). The mutational constraint spectrum quantified from variation in 141,456 humans. Nature 581, 434–443. 10.1038/s41586-020-2308-7.

30. Woodward Davis, A.S., Roozen, H.N., Dufort, M.J., DeBerg, H.A., Delaney, M.A., Mair, F., Erickson, J.R., Slichter, C.K., Berkson, J.D., Klock, A.M., et al. (2019). The human tissue-resident CCR5+ T cell compartment maintains protective and functional properties during inflammation. Sci. Transl. Med. 11. 10.1126/scitranslmed.aaw8718.

31. Stephens, J.C., Reich, D.E., Goldstein, D.B., Shin, H.D., Smith, M.W., Carrington, M., Winkler, C., Huttley, G.A., Allikmets, R., Schriml, L., et al. (1998). Dating the origin of the CCR5-Delta32 AIDS-resistance allele by the coalescence of haplotypes. Am. J. Hum. Genet. 62, 1507. 10.1086/301867.

32. Samson, M., Libert, F., Doranz, B.J., Rucker, J., Liesnard, C., Farber, M., Saragosti, S., Lapoumeroulie, C., Cognaux, J., Forceille, C., et al. (1996). Resistance to HIV-1 infection in Caucasian individuals bearing mutant alleles of the CCR-5 chemokine receptor gene. Nature 1996 382:6593 *382*, 722–725. 10.1038/382722a0.

33. Novembre, J., Galvani, A.P., and Slatkin, M. (2005). The Geographic Spread of the CCR5 Δ32 HIV-Resistance Allele. PLoS Biol. 3, e339. 10.1371/journal.pbio.0030339.

34. Ahsan, M., Ek, W.E., Rask-Andersen, M., Karlsson, T., Lind-Thomsen, A., Enroth, S., Gyllensten, U., and Johansson, Å. (2017). The relative contribution of DNA methylation and genetic variants on protein biomarkers for human diseases. PLoS Genet. 13, e1007005. 10.1371/journal.pgen.1007005.

35. Kurki, M.I., Karjalainen, J., Palta, P., Sipilä, T.P., Kristiansson, K., Donner, K.M., Reeve, M.P., Laivuori, H., Aavikko, M., Kaunisto, M.A., et al. (2023). FinnGen provides genetic insights from a well-phenotyped isolated population. Nature 2023 613:7944 *613*, 508–518. 10.1038/s41586-022-05473-8.

36. Wakefield, J. (2007). A Bayesian Measure of the Probability of False Discovery in Genetic Epidemiology Studies. Am. J. Hum. Genet. 81, 208. 10.1086/519024.

37. Roychoudhuri, R., Hirahara, K., Mousavi, K., Clever, D., Klebanoff, C.A., Bonelli, M., Sciumè, G., Zare, H., Vahedi, G., Dema, B., et al. (2013). Bach2 represses effector programmes to stabilize Treg-mediated immune homeostasis. Nature 498, 506. 10.1038/nature12199.

38. Muto, A., Tashiro, S., Nakajima, O., Hoshino, H., Takahashi, S., Sakoda, E., Ikebe, D., Yamamoto, M., and Igarashi, K. (2004). The transcriptional programme of antibody class switching involves the repressor Bach2. Nature 429, 566–571. 10.1038/nature02596.

39. Weinand, K., Sakaue, S., Nathan, A., Jonsson, A.H., Zhang, F., Watts, G.F.M., Al Suqri, M., Zhu, Z., Rao, D.A., Anolik, J.H., et al. (2024). The chromatin landscape of pathogenic transcriptional cell states in rheumatoid arthritis. Nature Communications 2024 15:1 *15*, 4650-. 10.1038/s41467-024-48620-7.

40. Ulirsch, J.C., Lareau, C.A., Bao, E.L., Ludwig, L.S., Guo, M.H., Benner, C., Satpathy, A.T., Kartha, V.K., Salem, R.M., Hirschhorn, J.N., et al. (2019). Interrogation of human hematopoiesis at single-cell and single-variant resolution. Nat. Genet. 51, 683. 10.1038/s41588-019-0362-6.

41. Rao, D.A., Gurish, M.F., Marshall, J.L., Slowikowski, K., Fonseka, C.Y., Liu, Y., Donlin, L.T., Henderson, L.A., Wei, K., Mizoguchi, F., et al. (2017). Pathologically expanded peripheral T helper cell subset drives B cells in rheumatoid arthritis. Nature 542, 110–114. 10.1038/nature20810.

42. Cuomo, A.S.E., Spenceley, E., Tanudisastro, H.A., Bowen, B., Henry, A., Huang, H.L., Xue, A., Zhou, W., Welland, M.J., Bryen, S.J., et al. (2025). Impact of rare and common genetic variation on cell type-specific gene expression in human blood. medRxiv 9, 2025.03.20.25324352. 10.1101/2025.03.20.25324352.

43. Giambartolomei, C., Vukcevic, D., Schadt, E.E., Franke, L., Hingorani, A.D., Wallace, C., and Plagnol, V. (2014). Bayesian Test for Colocalisation between Pairs of Genetic Association Studies Using Summary Statistics. PLoS Genet. 10, e1004383. 10.1371/JOURNAL.PGEN.1004383.

44. Wong, M.T.Y., Schölvinck, E.H., Lambeck, A.J.A., and Van Ravenswaaij-Arts, C.M.A. (2015). CHARGE syndrome: a review of the immunological aspects. European Journal of Human Genetics 23, 1451. 10.1038/ejhg.2015.7.

45. Stuart, P.E., Tsoi, L.C., Nair, R.P., Ghosh, M., Kabra, M., Shaiq, P.A., Raja, G.K., Qamar, R., Thelma, B.K., Patrick, M.T., et al. (2021). Transethnic analysis of psoriasis susceptibility in South Asians and Europeans enhances fine-mapping in the MHC and genomewide. HGG Adv. 3, 100069–100069. 10.1016/j.xhgg.2021.100069.

46. Mbatchou, J., Barnard, L., Backman, J., Marcketta, A., Kosmicki, J.A., Ziyatdinov, A., Benner, C., O’Dushlaine, C., Barber, M., Boutkov, B., et al. (2021). Computationally efficient whole-genome regression for quantitative and binary traits. Nat. Genet. 53, 1097–1103. 10.1038/s41588-021-00870-7.

47. Sakaue, S., Weinand, K., Isaac, S., Dey, K.K., Jagadeesh, K., Kanai, M., Watts, G.F.M., Zhu, Z., Brenner, M.B., McDavid, A., et al. (2024). Tissue-specific enhancer–gene maps from multimodal single-cell data identify causal disease alleles. Nature Genetics 2024 56:4 *56*, 615–626. 10.1038/s41588-024-01682-1.

48. Forde, S., Tye, B.J., Newey, S.E., Roubelakis, M., Smythe, J., McGuckin, C.P., Pettengell, R., and Watt, S.M. (2007). Endolyn (CD164) modulates the CXCL12-mediated migration of umbilical cord blood CD133+ cells. Blood 109, 1825–1833. 10.1182/blood-2006-05-023028.

49. Watt, S.M., Bühring, H.J., Simmons, P.J., and Zannettino, A.W.C. (2021). The stem cell revolution: on the role of CD164 as a human stem cell marker. NPJ Regen. Med. 6. 10.1038/s41536-021-00143-1.

50. Lagattuta, K.A., Park, H.L., Rumker, L., Ishigaki, K., Nathan, A., and Raychaudhuri, S. (2024). The genetic basis of autoimmunity seen through the lens of T cell functional traits. Nature Communications 2024 15:1 15, 1204-. 10.1038/s41467-024-45170-w.

51. Trynka, G., Sandor, C., Han, B., Xu, H., Stranger, B.E., Liu, X.S., and Raychaudhuri, S. (2013). Chromatin marks identify critical cell types for fine mapping complex trait variants. Nat. Genet. 45, 124–130. 10.1038/ng.2504.

52. Edwards, J.C.W., Szczepański, L., Szechiński, J., Filipowicz-Sosnowska, A., Emery, P., Close, D.R., Stevens, R.M., and Shaw, T. (2004). Efficacy of B-Cell–Targeted Therapy with Rituximab in Patients with Rheumatoid Arthritis. New England Journal of Medicine 350, 2572–2581. 10.1056/nejmoa032534.

53. Bucci, L., Hagen, M., Rothe, T., Raimondo, M.G., Fagni, F., Tur, C., Wirsching, A., Wacker, J., Wilhelm, A., Auger, J.P., et al. (2024). Bispecific T cell engager therapy for refractory rheumatoid arthritis. Nat. Med. 30, 1593–1601. 10.1038/s41591-024-02964-1.

54. Zhang, F., Jonsson, A.H., Nathan, A., Millard, N., Curtis, M., Xiao, Q., Gutierrez-Arcelus, M., Apruzzese, W., Watts, G.F.M., Weisenfeld, D., et al. (2023). Deconstruction of rheumatoid arthritis synovium defines inflammatory subtypes. Nature 2023 623:7987 623, 616–624. 10.1038/s41586-023-06708-y.

55. Farh, K.K.H., Marson, A., Zhu, J., Kleinewietfeld, M., Housley, W.J., Beik, S., Shoresh, N., Whitton, H., Ryan, R.J.H., Shishkin, A.A., et al. (2014). Genetic and epigenetic fine mapping of causal autoimmune disease variants. Nature 2014 518:7539 518, 337–343. 10.1038/nature13835.

56. Liu, Y.J., Miao, H.B., Lin, S., and Chen, Z. (2022). Association between rheumatoid arthritis and thyroid dysfunction: A meta-analysis and systematic review. Front. Endocrinol. (Lausanne). 13, 1015516. 10.3389/fendo.2022.1015516.

57. Raterman, H.G., Van Halm, V.P., Voskuyl, A.E., Simsek, S., Dijkmans, B.A.C., and Nurmohamed, M.T. (2008). Rheumatoid arthritis is associated with a high prevalence of hypothyroidism that amplifies its cardiovascular risk. Ann. Rheum. Dis. 67, 229–232. 10.1136/ard.2006.068130.

58. Guo, Y., Chung, W., Shan, Z., Zhu, Z., Costenbader, K.H., and Liang, L. (2023). Genome-Wide Assessment of Shared Genetic Architecture Between Rheumatoid Arthritis and Cardiovascular Diseases. J. Am. Heart Assoc. 12, 30211. 10.1161/JAHA.123.030211.

59. Sakaue, S., Gurajala, S., Curtis, M., Luo, Y., Choi, W., Ishigaki, K., Kang, J.B., Rumker, L., Deutsch, A.J., Schönherr, S., et al. (2023). Tutorial: a statistical genetics guide to identifying HLA alleles driving complex disease. Nature Protocols 2023 *10*, 1–17. 10.1038/s41596-023-00853-4.

60. Okada, Y., Kim, K., Han, B., Pillai, N.E., Ong, R.T.H., Saw, W.Y., Luo, M., Jiang, L., Yin, J., Bang, S.Y., et al. (2014). Risk for ACPA-positive rheumatoid arthritis is driven by shared HLA amino acid polymorphisms in Asian and European populations. Hum. Mol. Genet. 23, 6916–6926. 10.1093/hmg/ddu387.

61. Laufer, V.A., Tiwari, H.K., Reynolds, R.J., Danila, M.I., Wang, J., Edberg, J.C., Kimberly, R.P., Kottyan, L.C., Harley, J.B., Mikuls, T.R., et al. (2019). Genetic influences on susceptibility to rheumatoid arthritis in African-Americans. Hum. Mol. Genet. 28, 858–874. 10.1093/HMG/DDY395.

62. Traylor, M., Curtis, C., Patel, H., Breen, G., Lee, S.H., Xu, X., Newhouse, S., Dobson, R., Steer, S., Cope, A.P., et al. (2017). Genetic and environmental risk factors for rheumatoid arthritis in a UK African ancestry population: the GENRA case-control study. Rheumatology (Oxford) 56, 1282–1292. 10.1093/RHEUMATOLOGY/KEX048.

63. Verma, A., Damrauer, S.M., Naseer, N., Weaver, J.E., Kripke, C.M., Guare, L., Sirugo, G., Kember, R.L., Drivas, T.G., Dudek, S.M., et al. (2022). The Penn Medicine BioBank: Towards a Genomics-Enabled Learning Healthcare System to Accelerate Precision Medicine in a Diverse Population. J. Pers. Med. 12, 1974. 10.3390/jpm12121974.

64. Boutin, N.T., Schecter, S.B., Perez, E.F., Tchamitchian, N.S., Cerretani, X.R., Gainer, V.S., Lebo, M.S., Mahanta, L.M., Karlson, E.W., and Smoller, J.W. (2022). The Evolution of a Large Biobank at Mass General Brigham. J. Pers. Med. 12, 1323. 10.3390/jpm12081323.

65. Huang, S., Huang, J., Cai, T., Dahal, K.P., Cagan, A., He, Z., Stratton, J., Gorelik, I., Hong, C., Cai, T., et al. (2020). Impact of ICD10 and secular changes on electronic medical record rheumatoid arthritis algorithms. Rheumatology 59, 3759–3766. 10.1093/rheumatology/keaa198.

66. Hunter-Zinck, H., Shi, Y., Li, M., Gorman, B.R., Ji, S.G., Sun, N., Webster, T., Liem, A., Hsieh, P., Devineni, P., et al. (2020). Genotyping Array Design and Data Quality Control in the Million Veteran Program. Am. J. Hum. Genet. 106, 535–548. 10.1016/j.ajhg.2020.03.004.

67. Taliun, D., Harris, D.N., Kessler, M.D., Carlson, J., Szpiech, Z.A., Torres, R., Taliun, S.A.G., Corvelo, A., Gogarten, S.M., Kang, H.M., et al. (2021). Sequencing of 53,831 diverse genomes from the NHLBI TOPMed Program. Nature 590, 290–299. 10.1038/s41586-021-03205-y.

68. Mbatchou, J., Barnard, L., Backman, J., Marcketta, A., Kosmicki, J.A., Ziyatdinov, A., Benner, C., O’Dushlaine, C., Barber, M., Boutkov, B., et al. (2021). Computationally efficient whole-genome regression for quantitative and binary traits. Nat. Genet. 53, 1097–1103. 10.1038/s41588-021-00870-7.

69. Willer, C.J., Li, Y., and Abecasis, G.R. (2010). METAL: Fast and efficient meta-analysis of genomewide association scans. Bioinformatics 26, 2190–2191. 10.1093/bioinformatics/btq340.

70. Maller, J.B., McVean, G., Byrnes, J., Vukcevic, D., Palin, K., Su, Z., Howson, J.M.M., Auton, A., Myers, S., Morris, A., et al. (2012). Bayesian refinement of association signals for 14 loci in 3 common diseases. Nat. Genet. 44, 1294–1301. 10.1038/NG.2435.

71. Ma, S., Zhang, B., LaFave, L.M., Earl, A.S., Chiang, Z., Hu, Y., Ding, J., Brack, A., Kartha, V.K., Tay, T., et al. (2020). Chromatin Potential Identified by Shared Single-Cell Profiling of RNA and Chromatin. Cell 183, 1103–1116.e20. 10.1016/J.CELL.2020.09.056.

72. Mimitou, E.P., Lareau, C.A., Chen, K.Y., Zorzetto-Fernandes, A.L., Hao, Y., Takeshima, Y., Luo, W., Huang, T.S., Yeung, B.Z., Papalexi, E., et al. (2021). Scalable, multimodal profiling of chromatin accessibility, gene expression and protein levels in single cells. Nat. Biotechnol. 39, 1246–1258. 10.1038/S41587-021-00927-2.

73. Chen, A.F., Parks, B., Kathiria, A.S., Ober-Reynolds, B., Goronzy, J.J., and Greenleaf, W.J. (2022). NEAT-seq: simultaneous profiling of intra-nuclear proteins, chromatin accessibility and gene expression in single cells. Nature Methods 2022 19:5 19, 547–553. 10.1038/s41592-022-01461-y.

74. Luecken, M.D., Burkhardt, D.B., Cannoodt, R., Lance, C., Agrawal, A., Aliee, H., Chen, A.T., Deconinck, L., Detweiler, A.M., Granados, A., et al. (2021). A sandbox for prediction and integration of DNA, RNA, and proteins in single cells. Proceedings of the Neural Information Processing Systems Track on Datasets and Benchmarks 1.

75. Jia, X., Han, B., Onengut-Gumuscu, S., Chen, W.M., Concannon, P.J., Rich, S.S., Raychaudhuri, S., and de Bakker, P.I.W. (2013). Imputing Amino Acid Polymorphisms in Human Leukocyte Antigens. PLoS One 8, e64683. 10.1371/journal.pone.0064683.

76. Das, S., Forer, L., Schönherr, S., Sidore, C., Locke, A.E., Kwong, A., Vrieze, S.I., Chew, E.Y., Levy, S., McGue, M., et al. (2016). Next-generation genotype imputation service and methods. Nat. Genet. 48, 1284–1287. 10.1038/ng.3656.

