## Supplementary Figures and Text for "Multi-ancestral GWAS with the VA Million Veteran Program enables functional interpretation of rheumatoid arthritis alleles"

Sakaue et al.

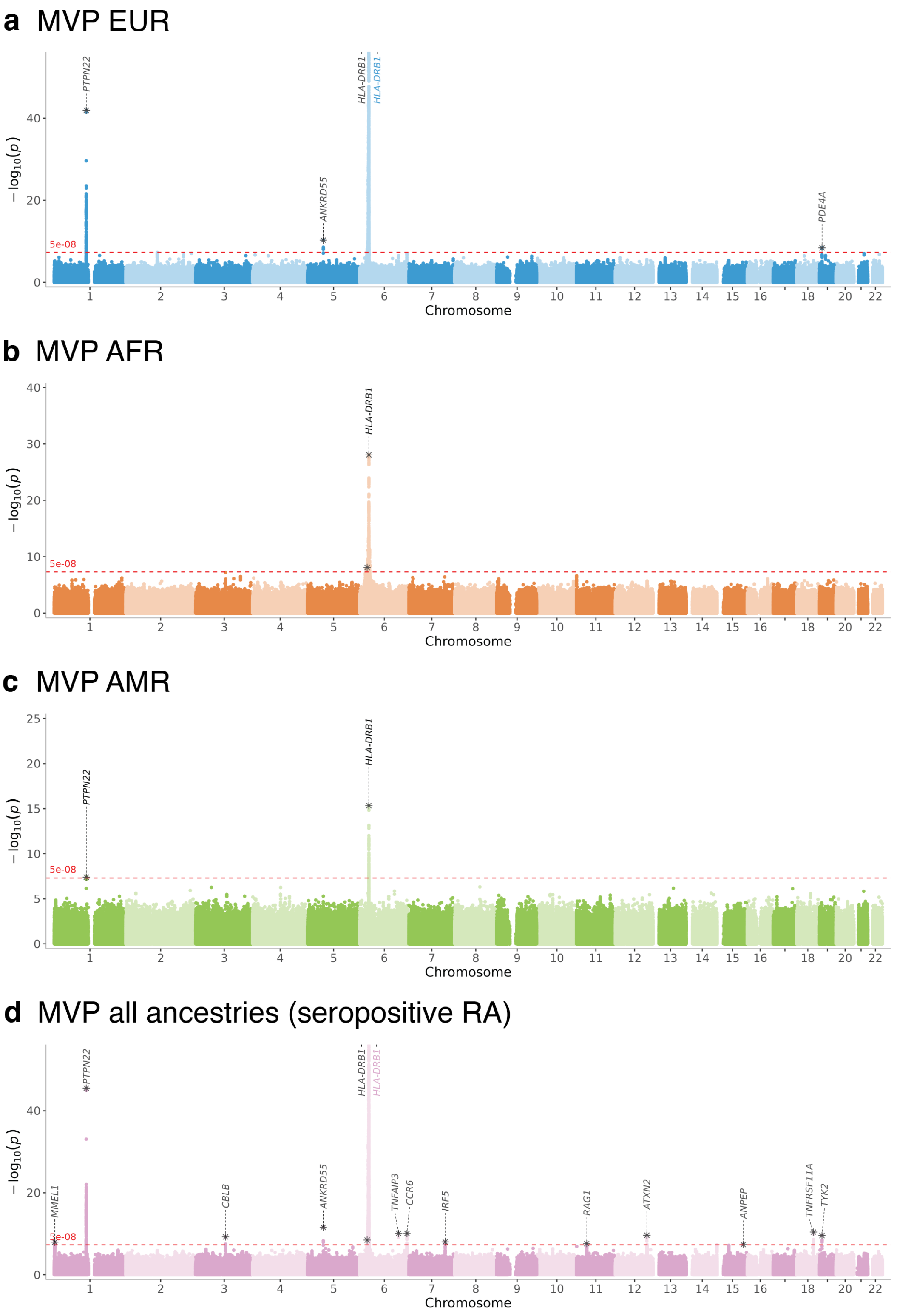

### Supplementary Figure 1 I Manhattan plots of MVP GWAS.

Manhattan plots of MVP RA GWAS in EUR (a), AFR (b), and AMR (c) ancestries, and MVP-wide association for seropositive RA (d). Lead variants with asterisks are annotated by the closest genes.

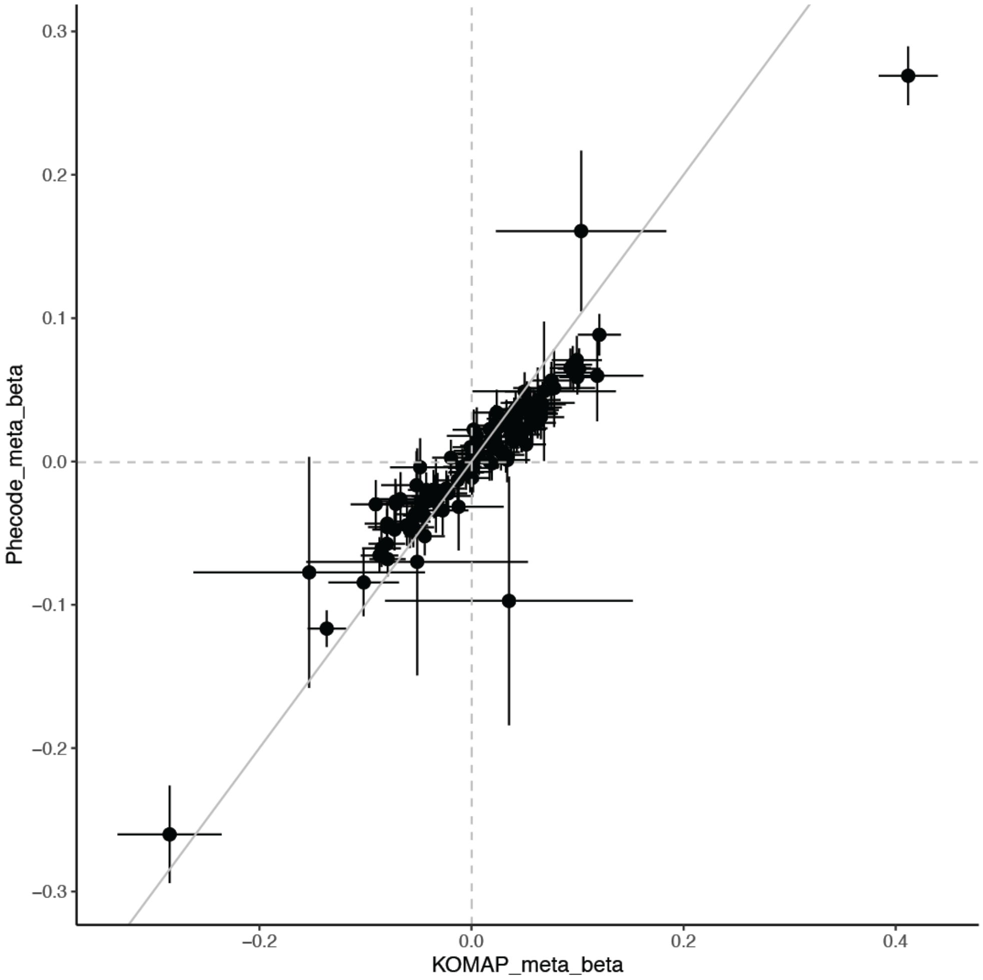

Supplementary Figure 2 I Effect size comparison between KOMAP RA phenotyping and phecode using ICD10.

We plotted the effect sizes (beta) of RA of previously identified 109 autosomal loci from Ishigaki et al. between our MVP GWAS using KOMAP RA phenotype (x-axis) and the MVP GWAS using phecode using ≥2 RA ICD codes (y-axis).

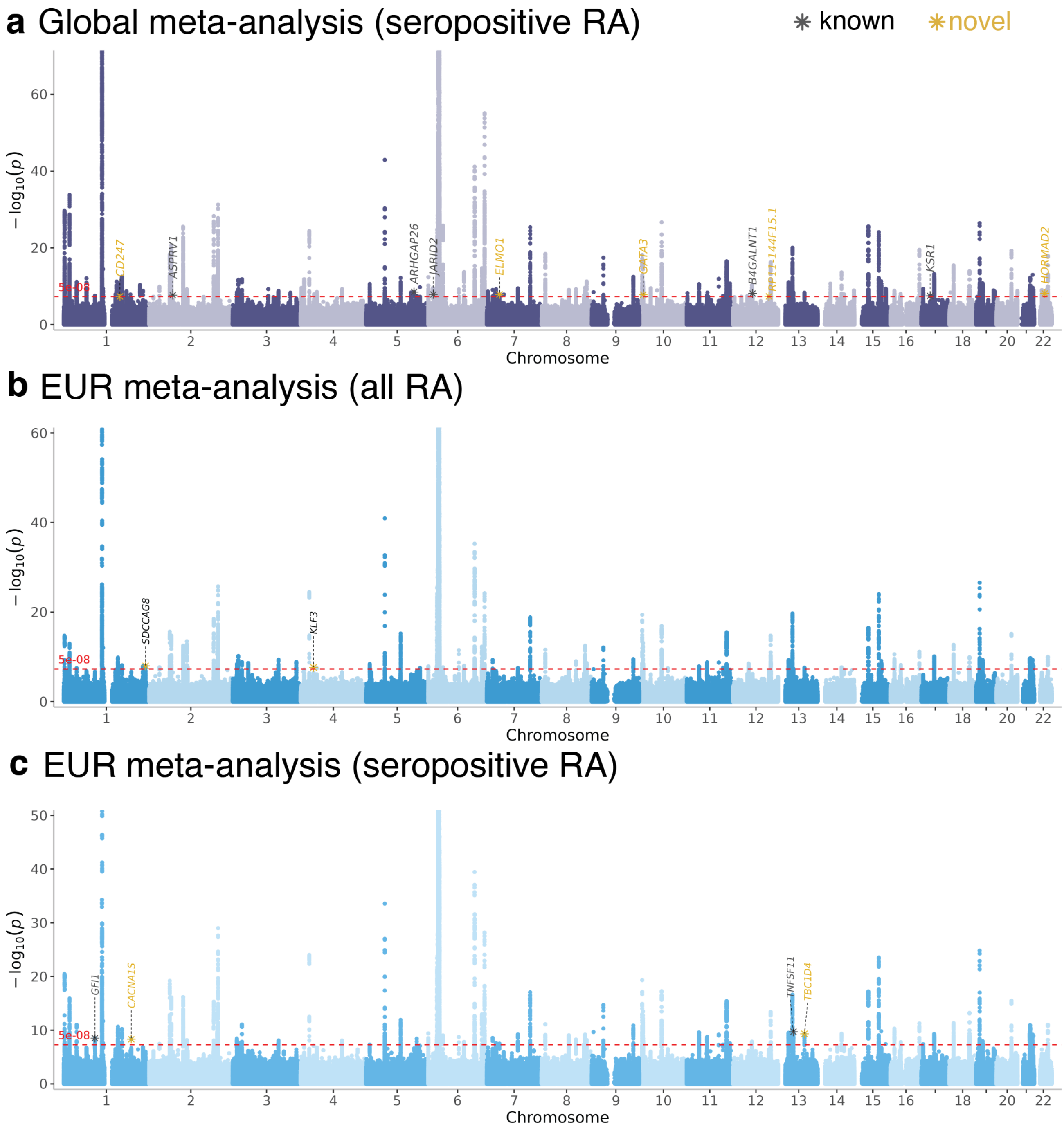

**Supplementary Figure 3 | Manhattan plots of meta-analyzed RA cohorts with MVP.** Manhattan plots of joint meta-analyses between MVP and all previous RACI cohorts in seropositive RA (a), EUR-only (b), and seropositive RA in EUR-only (c). Lead variants with asterisks are annotated by the closest genes and colored in grey (previously reported loci) or in yellow (novel loci).

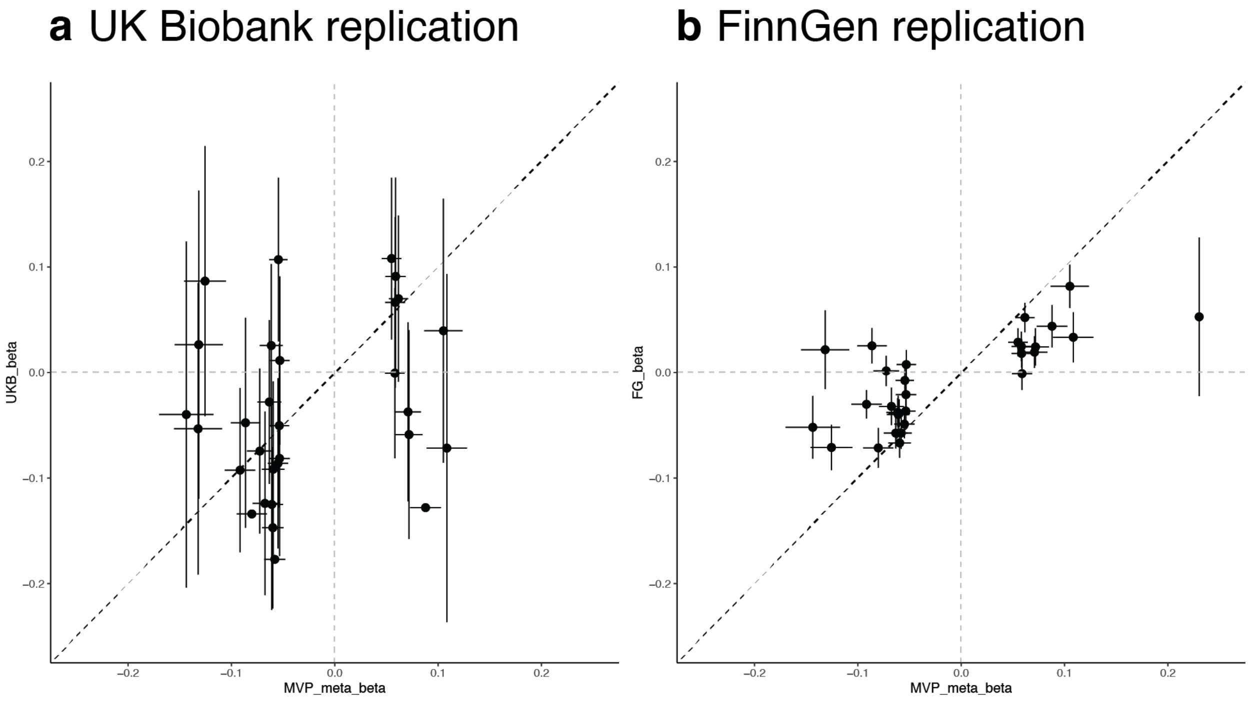

### Supplementary Figure 4 I Replication with UK Biobank and FinnGen.

We plotted concordance of effect sizes (beta) from our novel loci between our MVP study (x-axis) and UK Biobank (a) or FinnGen (b) (y-axis). Bars indicate standard errors.

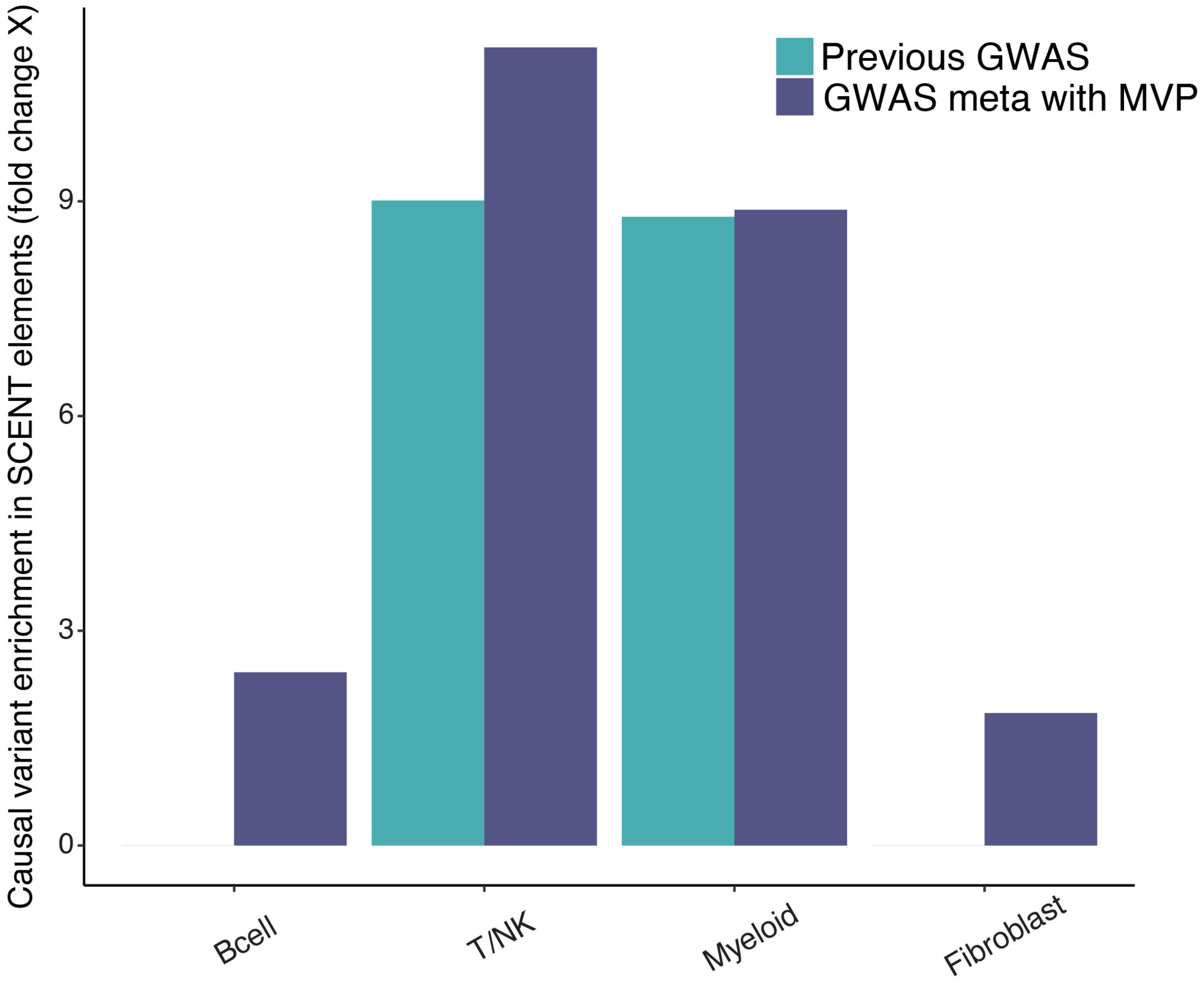

Supplementary Figure 5 I Enrichment of GWAS causal variants within cell-type-specific functional regulatory elements by SCENT.

Causal variant enrichment for RA GWAS within cell-type-specific SCENT regulatory elements. Cell types are indicated on the x-axis, and bars are colored based on previous Ishigaki et al. GWAS (green) or our updated GWAS meta-analysis with MVP (purple).

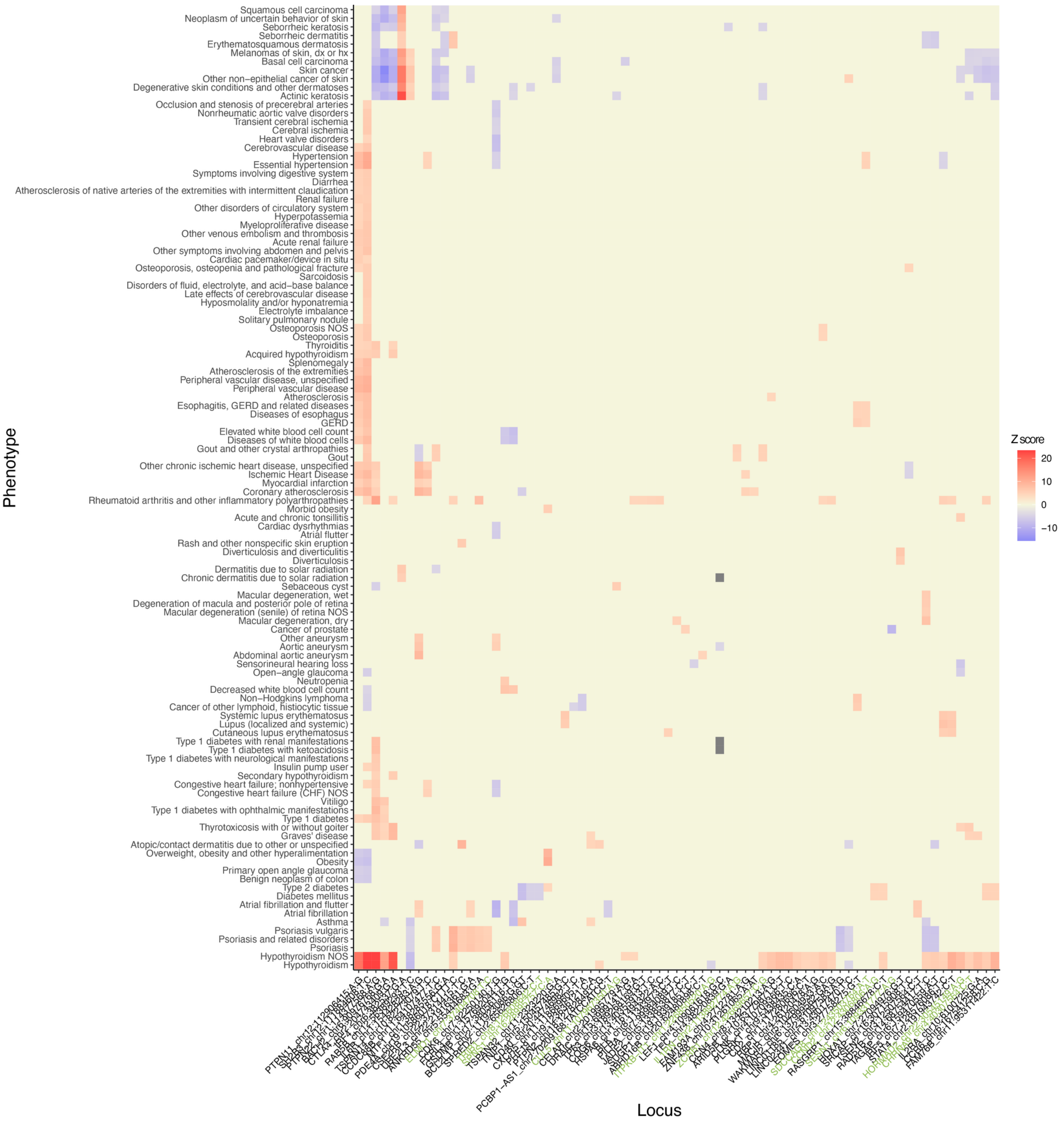

### Supplementary Figure 6 | Phenotypic characterization of RA loci.

MVP PheWAS of RA GWAS loci. The sign of Z scores represent the effect direction compared with RA alleles with positive Z scores being consistent with RA effect direction and negative Z scores being opposite from RA effect direction.

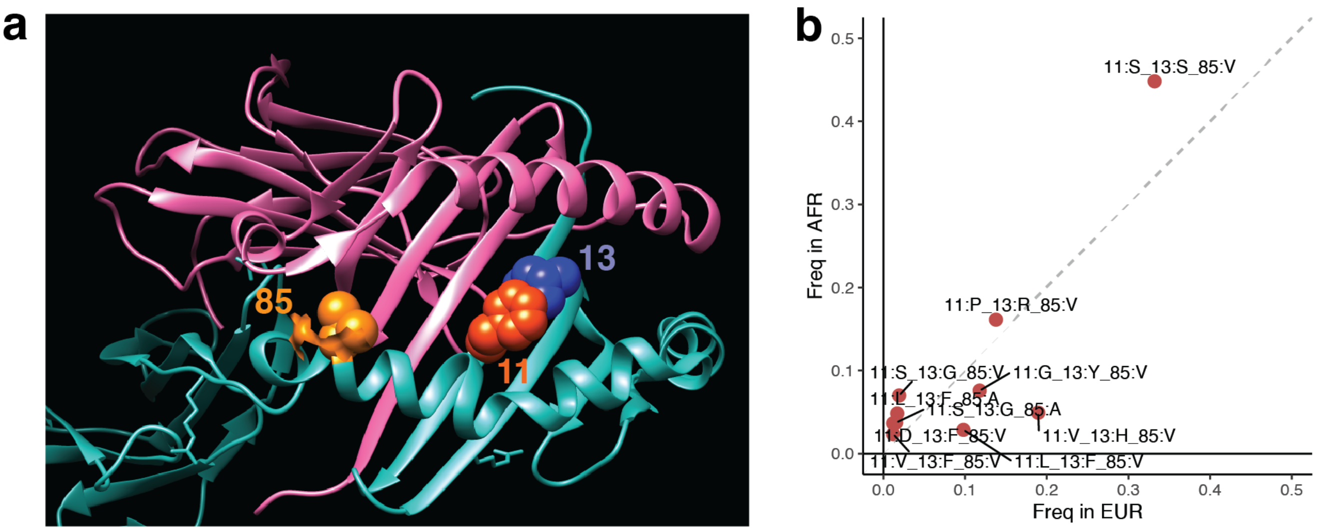

Supplementary Figure 7 l HLA associations at MVP.

a. Positional information of the independent association signal of HLA-DRB1 amino acid residues in MVP. This figure was prepared with UCSF chimera based on Protein Data Bank entries 3pdob. b. Allele frequency differences between EUR (x-axis) and AFR (y-axis) of HLA haplotypes consisted of independent amino acid positions of RA risk identified from AFR.

### Supplementary Text | A full list of VA Million Veteran Program researchers

**VA Million Veteran Program (MVP)**

*MVP Program Office*

- Sumitra Muralidhar, Ph.D., Program Director
  US Department of Veterans Affairs, 810 Vermont Avenue NW, Washington, DC 20420
- Jennifer Moser, Ph.D., Associate Director, Scientific Programs
  US Department of Veterans Affairs, 810 Vermont Avenue NW, Washington, DC 20420
- Jennifer E. Deen, B.S., Associate Director, Cohort & Public Relations
  US Department of Veterans Affairs, 810 Vermont Avenue NW, Washington, DC 20420

*MVP Steering Committee*

- Co-Chair: Philip S. Tsao, Ph.D.
  VA Palo Alto Health Care System, 3801 Miranda Avenue, Palo Alto, CA 94304
- Co-Chair: Sumitra Muralidhar, Ph.D.
  US Department of Veterans Affairs, 810 Vermont Avenue NW, Washington, DC 20420
- J. Michael Gaziano, M.D., M.P.H.
  VA Boston Healthcare System, 150 S. Huntington Avenue, Boston, MA 02130
- Adriana Hung, M.D., M.P.H.,
  VA Tennessee Valley Healthcare System, 1310 24th Avenue, South Nashville, TN 37212
- Dave Oslin, M.D.
  Philadelphia VA Medical Center, 3900 Woodland Avenue, Philadelphia, PA 19104
- Deepak Voora, M.D.
  Durham VA Medical Center, 508 Fulton Street, Durham, NC 27705

*MVP Co-Principal Investigators*

- J. Michael Gaziano, M.D., M.P.H.
  VA Boston Healthcare System, 150 S. Huntington Avenue, Boston, MA 02130

- Philip S. Tsao, Ph.D.
  VA Palo Alto Health Care System, 3801 Miranda Avenue, Palo Alto, CA 94304

*MVP Core Operations*

- Jessica V. Brewer, M.P.H., Director, MVP Cohort Operations
  VA Boston Healthcare System, 150 S. Huntington Avenue, Boston, MA 02130
- Mary T. Brophy M.D., M.P.H., Director, VA Central Biorepository
  VA Boston Healthcare System, 150 S. Huntington Avenue, Boston, MA 02130
- Kelly Cho, M.P.H, Ph.D., Director, MVP Phenomics
  VA Boston Healthcare System, 150 S. Huntington Avenue, Boston, MA 02130
- Lori Churby, B.S., Director, MVP Regulatory Affairs
  VA Palo Alto Health Care System, 3801 Miranda Avenue, Palo Alto, CA 94304
- Jacob T. Kean, Ph.D., Acting Director, VA Informatics and Computing Infrastructure (VINCI)
  VA Salt Lake City Health Care System, 500 Foothill Drive, Salt Lake City, UT 84148
- Saiju Pyarajan Ph.D., Director, Data and Computational Sciences
  VA Boston Healthcare System, 150 S. Huntington Avenue, Boston, MA 02130
- Robert Ringer, Pharm.D., Director, VA Albuquerque Central Biorepository
  New Mexico VA Health Care System, 1501 San Pedro Drive SE, Albuquerque, NM 87108
- Luis E. Selva, Ph.D., Director, MVP Biorepository Coordination
  VA Boston Healthcare System, 150 S. Huntington Avenue, Boston, MA 02130
- Shahpoor (Alex) Shayan, M.S., Director, MVP PRE Informatics
  VA Boston Healthcare System, 150 S. Huntington Avenue, Boston, MA 02130
- Brady Stephens, M.S., Principal Investigator, MVP Information Center
  Canandaigua VA Medical Center, 400 Fort Hill Avenue, Canandaigua, NY 14424
- Stacey B. Whitbourne, Ph.D., Director, MVP Cohort Development and Management
  VA Boston Healthcare System, 150 S. Huntington Avenue, Boston, MA 02130
